# As rates of ASD and ADHD rise, genetic contributions fall: Evidence for widening diagnostic criteria

**DOI:** 10.1101/2025.11.10.25339891

**Authors:** Sonja LaBianca, Mette Lise Lousdal, Morten Dybdahl Krebs, Ole S Hansen, Kajsa-Lotta LA Georgii Hellberg, Mischa Lundberg, Johanne Østerby Sørensen, Jesper R Gaadin, Henrik Ohlsson, iPSYCH Study Consortium, Anders Borglum, Esben Agerbo, Thomas Werge, Clara Albinana, Bjarni J Vilhjalmsson, Kenneth S Kendler, Oleguer Plana-Ripoll, Andrew J Schork

## Abstract

**Importance:** The incidence of ADHD and autism spectrum disorder (ASD) has increased markedly over recent decades, raising concerns about the emergence of new risk factors. Current literature typically attributes increased rates to changes in diagnostic practice, stigmatization, and awareness, but critically few studies have explored changes in underlying risk factors.

**Objective:** To assess changes in the genetic risk profile of individuals diagnosed with ASD or ADHD according to year of incident diagnosis.

**Design:** We used the iPSYCH2015 study, a population-based case-cohort with complete ascertainment of incident diagnoses for ASD and ADHD made from 1994 to 2016.

**Setting:** Denmark.

**Participants:** ASD (N=17,071) and ADHD (N=20,111).

**Exposure:** Year of incident diagnosis. Regression models tested changes in the mean genetic risk profile of individuals diagnosed in each consecutive year (1994-2016), adjusting for age, sex, and ancestry.

**Main Outcomes:** We used polygenic scores for psychiatric (ADHD, ASD, depression, bipolar, schizophrenia) and cognitive-behavioral (addiction, educational attainment, IQ, neuroticism, risk-taking) outcomes to capture the genetic risk profiles of diagnosed individuals.

**Results:** A more recent ADHD diagnosis was associated (p<0.001) with less genetic risk for ADHD (β=-0.06 SD per 10 years) and other disorders (ASD, bipolar, schizophrenia). Similarly, a more recent ASD diagnosis was associated with less genetic risk for ASD (β=-0.07) and other disorders/traits (bipolar, schizophrenia, educational attainment).

**Conclusions and Relevance:** Our novel approach suggests that over recent decades diagnostic practice around ADHD and ASD has evolved to capture a different profile of genetic risk. These findings support broadening diagnostic criteria as the explanation for the rise in incidence, with implications for understanding prevalence trends in relation to changes in risk factors and clinical practice.

**KEY POINTS:** *Question:* Has the genetic risk profile of individuals diagnosed with autism spectrum disorder (ASD) and ADHD changed as diagnostic rates have risen?

*Findings:* In the iPSYCH case-cohort study, we observed the genetic contributions to ASD and ADHD diagnoses have weakened over the past two decades.

*Meaning:* Our results suggest that recent increase in ASD and ADHD diagnoses coincide with a broadening of diagnostic criteria.

## INTRODUCTION

Over recent decades rates of ADHD and autism spectrum disorder (ASD) diagnoses have increased remarkably, with reports from four-to ten-fold across multiple populations^1–4^. This has sparked debates regarding the emergence of novel environmental risk factors and impacts of changing clinical practice or societal awareness^5^. The nosology of ASD and ADHD has evolved significantly over the last 40 years^6–8^. Initially conceived as distinct, narrowly defined childhood disorders, they are now considered life-long conditions with a spectrum of manifestations and high rates of co-occurring mental health difficulties^9,10^. Additionally, there has been increased clinical and public awareness driven in part by concerns of underdiagnosis^9,11^. Undiagnosed ADHD, for example, is associated with negative outcomes such as unemployment, criminality, and premature mortality^11,12^. Conversely, sharp increases in diagnostic rates has brought about concerns of overdiagnosis^13–17^, which can lead to unnecessary treatments and adverse side-effects of interventions^18^, or be a consequence of social services requiring a diagnosis to allocate support^15,17^. Furthermore, changes in the environment have been proposed, both in terms of novel exposures (e.g., perinatal acetaminophen exposure^19^ or screen time^20^) but also societal changes (e.g., school environment^21^). Finally, the increased emphasis on diagnosis in females, adolescents, and adults^2,3^, where clinical manifestations differ from previously predominant childhood, male presentations^10,22–24^, further complicates discussions. Although increasing diagnostic rates are often attributed to the interplay of these changes^9,11^, few have evaluated if underlying risk factors are also changing.

Both ADHD and ASD are highly heritable disorders (∼80%)^25,26^ indicating a large etiological contribution from genes. Environmental exposures, clinical guidelines, and societal factors evolve continuously or may change abruptly. An individual’s inherited genetic risk, however, remains fixed throughout their lifespan and population levels are constant over recent decades^27^. This creates a unique opportunity to study the relationship between time-varying diagnostic trends and time-invariant genetic factors to explore if diagnoses evolve to capture different constellations of risk. Only two studies have examined this relationship for ASD^28^ and ADHD^29^ using indirect measures of genetics and could not detect evidence of changing genetic or environmental risk contributions. Our recent work^30^ was the first to use polygenic risk score (PGS), emerging tools for directly indexing the inherited genetic risk carried by an individual that offer promise in clinical and etiological studies^31^. We showed that individuals diagnosed with any of multiple psychiatric disorders, including ASD and ADHD, carried lower PGS if born (and diagnosed) more recently^27^. However, in this study, the PGS were restricted to those of the index disorder, and did not explore how the genetic risk for other disorders/traits changed over time. Taken together the limited studies and current relevance of changing rates of ADHD and ASD necessitate deeper, targeted investigations and emerging studies^32^ suggest more powerful applications of PGS (i.e., using multiple scores to disambiguate among explanations) could add clarity.

In this study, we take a novel approach to examine and disambiguate among potential explanations for temporal changes in the genetic contributions to ADHD and ASD. We show that an analysis of multiple PGS (genetic risk profile) can clarify among potential drivers of increasing diagnostic rates: 1) a lowering threshold, consistent with less severe symptomatology or emerging environmental risk factors, 2) shifting diagnostic boundaries, allowing for different clinical manifestations previously assigned to other diagnoses, or 3) better detection, which implicates catching up on previously undiagnosed individuals. We use the iPSYCH2015 case-cohort study^33,34^, the largest genetic research cohort of ADHD and ASD with diagnoses spanning over two decades, to test how associated genetic risk profiles are changing over time and assess their consistency with these potential explanations.

## METHOD

### Study population

The Lundbeck Foundation initiative for Integrative Psychiatric Research (iPSYCH2015)^33,34^ is a Danish case-cohort study using national biobanks and registers. The source population consists of all singleton births May 1, 1981 and December 31, 2008, alive and residing in Denmark at age one-year-old (N=1,657,449). The sample (N=143,265) selected all incident diagnoses (N=93,608) of major psychiatric disorders through 2015 and a random population cohort (N=50,615). This study included individuals with a registered diagnosis of ADHD (N=29,668; ICD-10: F90.0) and/or ASD (N=24,975; ICD-10: F84.0, F84.1, F84.5, F84.8 or F84.9) in the Danish Psychiatric Central Research Register (PCR)^35^ between January 1, 1994, and December 31, 2015. Use of iPSYCH2015 data follows Danish Scientific Ethics Committee, the Danish Health Data Authority, the Danish Data Protection Agency, and the Danish Neonatal Screening Biobank Steering Committee. Data access was via secure portals in accordance with guidelines set by the Danish Data Protection Agency, the Danish Health Data Authority, and Statistics Denmark.

Diagnostic rates for ADHD and ASD were displayed in two-dimensional histograms using R package *ggplot2*, presenting 1×1-year bins with year of first diagnosis (year) along the *x*-axis, and age at first diagnosis (age) along the *y*-axis. Color coding indicates count of diagnoses in each bin restricting to >4 individuals to ensure data protection.

### Genotyping and quality controls

DNA was extracted and amplified from dried blood spots at the Danish Neonatal Screening Biobank^36^ and genotyped using the Illumina Global Screening or the Infinium PsychChip v1.0 Array. Data quality control is described elsewhere^33,34^. Imputation followed the RICOPILI pipeline^37^ using the Haplotype Reference Consortium panel^38^. Ancestry principal component (PC) analysis was conducted to create the largest homogenous group for analysis, a sample of broadly northern European genetic ancestry, and retain 10 PCs as analytical covariates.

### Genetic risk profiles

Genetic risk profiles included 10 PGS constructed using results from genome wide association studies (GWAS) in public repositories. GWAS include five psychiatric disorders; ADHD, ASD, Bipolar disorder (BP), Major Depressive disorder (MDD), and Schizophrenia (SCZ) and five cognitive/behavioral traits; Educational Attainment (EA), Intelligence Quotient (IQ), Risk-taking, Neuroticism and Addiction PGS (Supplementary Table 1). Meta-PRS^39^ was used to compute PGS from five psychiatric disorders, which improves prediction by combining external GWAS data and internal individual-level genetic data. For the cognitive/behavioral traits, where internal GWAS was not available, PGS were computed with LDpred2^40^.

### Statistical Analysis

Linear regressions were fitted with each PGS scaled to the population cohort as the outcome and year, age, sex as recorded in the registers, and 10 PCs as predictors. We adjusted for age to account for increasing age at diagnosis in more recent years^3^. Year trends for PGS were depicted by projecting the expected PGS from the fitted model over a range of years, holding other covariables constant at their mean. Post-hoc sensitivity analyses explored the robustness of results: First, since age and year are confounded in a birth cohort^41^, the goodness of fit and consistency of effect sizes for year adjusted by age and other covariates was compared to alternative models including: covariates only, covariates and year, covariates and age, covariates, age and birth cohort, or covariates and birth cohort. Second, non-linear effects of year were explored by adding quadratic terms and comparing the fit to the linear only models. Sex differences were tested by including interaction terms (sex by year) and comparing the model fit. Finally, the effects of year on meta-PRS^39^ were compared to those on PGS computed with LDpred2^40^ using only external GWAS. Significance was determined by Bonferroni adjustment for two disorders and 10 PGS (p<0.05/(2*10)=0.0025). All analyses were conducted using Rv4.1.3 (R Core Team 2022).

## RESULTS

### Diagnostic rates of ADHD and ASD increase with time

Previous reports have shown an increasing diagnostic rate of ASD and ADHD^3^. Here we depict these trends in our specific analysis samples. From the iPSYCH case-cohort study, we include 20,111 individuals diagnosed with ADHD first diagnosed between January 1^st^ 1994 to December 31^st^ 2015 (Figure 1A, Supplementary Table 2) and 17,071 individuals first diagnosed with ASD in the same period (Figure 1B, Supplementary Table 3). We see, for both disorders, an appreciable increase in the number of diagnoses at a given age with increasing year (increasing brightness from left to right, Figure 1).

**Figure 1.**
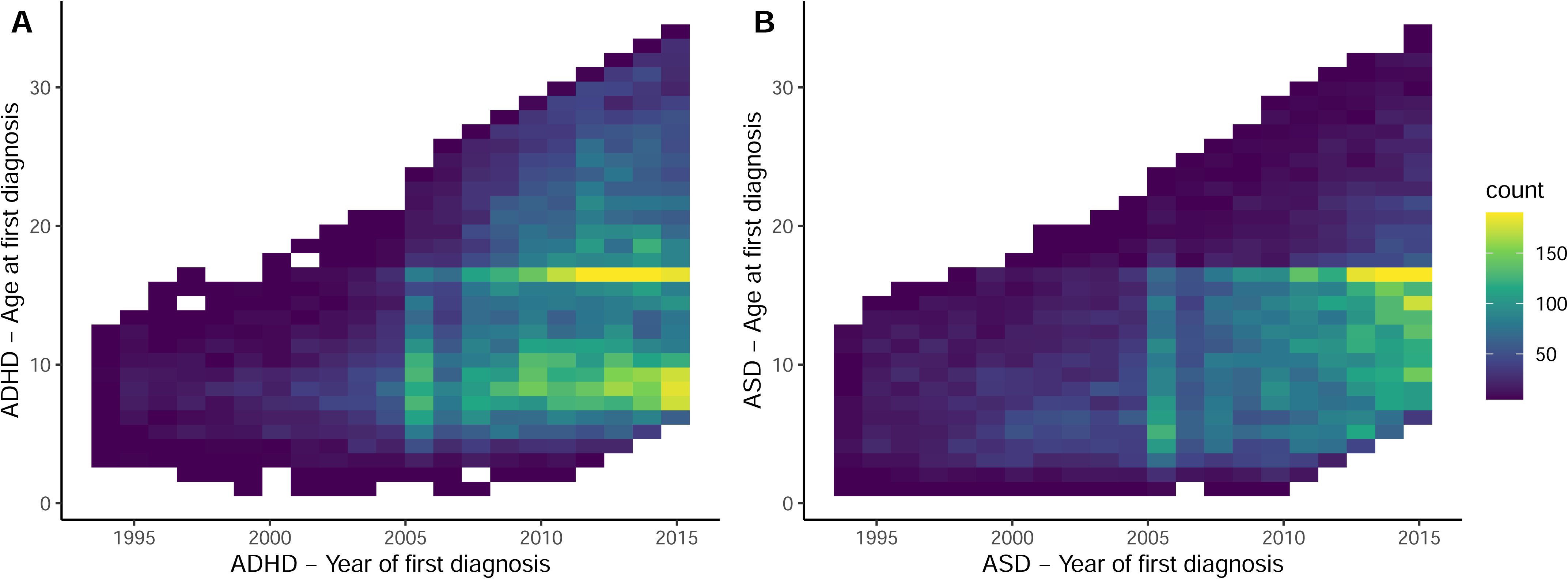
A) ADHD population N=20,111 and **B)** ASD population N=17,071, respectively, with distribution of year (year of diagnosis) on the x-axis and age (age of diagnosis) on the y-axis. Each cell represents counts of individuals diagnosed in that particular year and age, due to data protection restricting to cells >4 individuals.

### Genetic risk profiles can clarify mechanisms of increasing diagnostic rates

We conducted simulations to frame expectations for how genetic risk profiles are impacted by different processes of increasing rates (Figure 2, Supplementary Note). Our model posits new cases are diagnosed from one of three undiagnosed segments of the populations: 1. individuals from the general population with milder forms of the index disorder (referred to as “lowering threshold”), and 2. individuals with a set of symptoms consistent with or previously referred to as a second disorder (referred to as “shifting boundaries”). 3. individuals with a set of symptoms consistent with the index disorder but who were not previously diagnosed (referred to as “better detection”). Under a lowering threshold, we expect the index PGS to be reduced and secondary disorder PGS to change according to genetic correlation, in this scenario be reduced. For shifting boundaries, we expect the index disorder PGS to similarly decrease, but PGS for the secondary disorders to increase, regardless of genetic correlation. For better detection, we expect increases in the differences in PGS between cases and controls, with an increase of secondary scores in proportion to genetic correlation. These vignettes are confirmed with simulation experiments (Supplementary Note).

**Figure 2.**
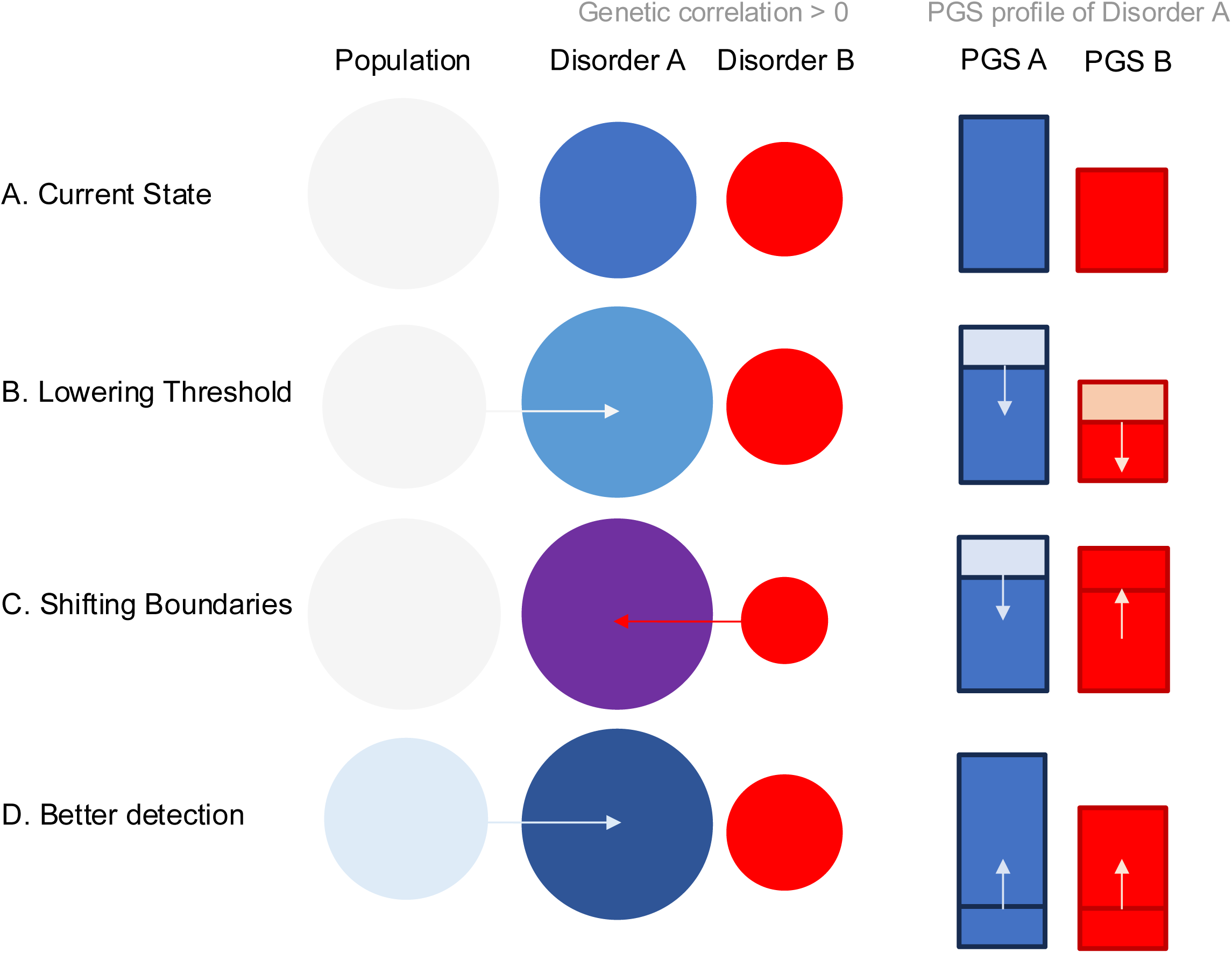
Genetic risk profiles are expected to be impacted differently under different scenarios of increasing diagnoses. Here, a profile of two polygenic scores (PGS) is considered, one for an index disorder (A, blue) and one for a second disorder (B, red) that is assumed to share genetic contributions with the index disorder (genetic correlation, r_G_, > 0). The expected profile for a current case with disorder A has high levels of PGS disorder A and modest levels for disorder B due to the r_G_ (A). If the number with disorder A is expanded, by a lowering threshold both PGS A and PGS B are expected to decrease (B). If expansion is due to shifting boundaries, PGS A is expected to decrease, while PGS B would increase. If expansion is due to better detection, PGS A and PGS B will appear to increase because the controls are expected to decrease (D).

### ADHD diagnoses associate with less genetic risk in more recent years

Regression models (Figure 3, Table 1, Supplementary Tables 4-13) suggested the mean PGS of ADHD cases decreased significantly with increasing year (β=-0.06 SD per 10 years; p=0.001), but not with increasing age (β<0.01, p=0.81). The mean level of other PGS also decreased with increasing year: ASD (β=-0.07, p<0.001), BP (β=-0.06, p=0.001) and SCZ (β=-0.06, p<0.001). The effects of age were inconsistent across the PGS; while MDD, BP, SCZ, Addiction and Neuroticism significantly increased, ASD, EA and IQ decreased (Supplementary Figure 1, Supplementary Tables 4-13). These results appear consistent with a lowering threshold for increasing year of ADHD diagnosis (Figure 2-3)

**Figure 3.**
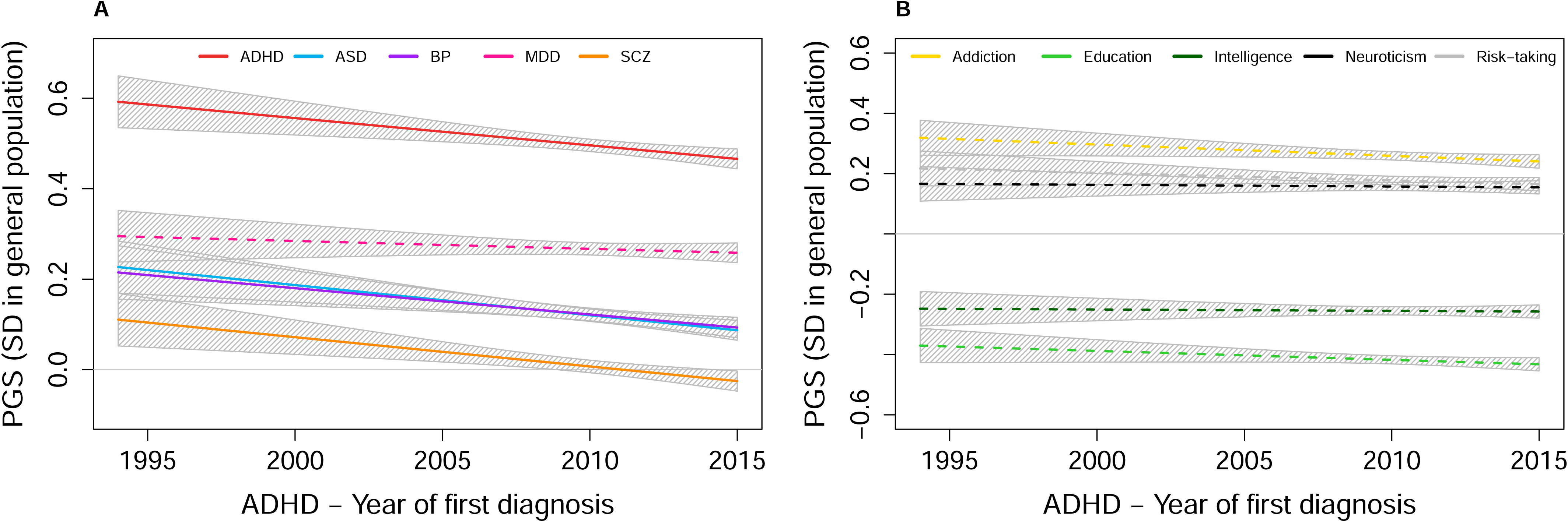
Genetic risk profile in ADHD patients (N=20,111) reveals decreasing trends for year of diagnosis between 1994:2015 on x-axis. The y-axis shows polygenic score (PGS) level in standard deviations (SD) to the general population. Each line represents the regression results from each of the five psychiatric PGS in **A)** and five cognitive/behavioral PGS in **B)** predicted by year while adjusting for age and covariate, shown in **Table 1**. After Bonferroni correction (<0.05/20) solid lines are significant and dashed lines are not.

**Table 1.** Linear regression predicting polygenic scores from year of diagnosis adjusted by age and covariates reveals a decreasing profile of genetic risk in ADHD (N=20,111) and ASD (N=17,071).

| Polygenic scores | Year of ADHD diagnosis | Year of ASD diagnosis |
| --- | --- | --- |
| ADHD | -0.06 (-0.09:-0.03), p=0.001** | 0.04 (0.01:0.07), p=0.005* |
| ASD | -0.07 (-0.10:-0.03), p<0.001** | -0.07 (-0.10:-0.04), p<0.001** |
| BP | -0.06 (-0.09:-0.02), p=0.001** | -0.05 (-0.08:-0.02), p=0.001** |
| MDD | -0.02 (-0.05:0.01), p=0.321 | 0.01 (-0.02:0.04), p=0.655 |
| SCZ | -0.06 (-0.10:-0.03), p<0.001** | -0.05 (-0.08:-0.02), p=0.001** |
| Addiction | -0.04 (-0.07:-0.00), p=0.037* | -0.02 (-0.05:0.01), p=0.257 |
| Education | -0.03 (-0.06:0.00), p=0.093 | -0.10 (-0.13:-0.07), p<0.001** |
| Intelligence | -0.00 (-0.04:0.03), p=0.801 | -0.03 (-0.07:-0.00), p=0.030* |
| Risk-taking | -0.02 (-0.06:0.01), p=0.167 | -0.04 (-0.07:-0.00), p=0.026* |
| Neuroticism | -0.01 (-0.04:0.03), p=0.752 | 0.01 (-0.02:0.04), p=0.378 |

Year=year of an individual's first recorded diagnosis, Age=age at an individual's first recorded diagnosis, Covariates=sex and 10 principal components of ancestry, ADHD=attention deficit hyperactivity disorder, ASD=autism spectrum disorder, BP=bipolar disorder, MDD=major depressive disorder, SCZ=schizophrenia spectrum disorder. Significance <0.05\* and <0.05/20\*\*.

### ASD diagnoses associate with less genetic risk in more recent years

Regression models in ASD (Figure 4, Table 1, Supplementary Tables 14-23) also suggested the ASD PGS significantly decreased with increasing year (β=-0.07 SD per 10 years, p<0.001), but not age (β=-0.01, p=0.460). With increasing year, we observed significant decreases in other PGS: BP (β=-0.05, p=0.001), SCZ (β=-0.05, p=0.001) and EA (β=-0.10, p=<0.001). We observed a nominally significant increase in ADHD PGS (β=0.04, p=0.005). The effects of age in ASD patients were inconsistent across PGS; while MDD, Addiction and Neuroticism significantly increased, EA decreased (Supplementary Figure 2, Supplementary Tables 14-23). For ASD our results also appear consistent with a lowering threshold with increasing year (Figure 2, Figure 4)

**Figure 4.**
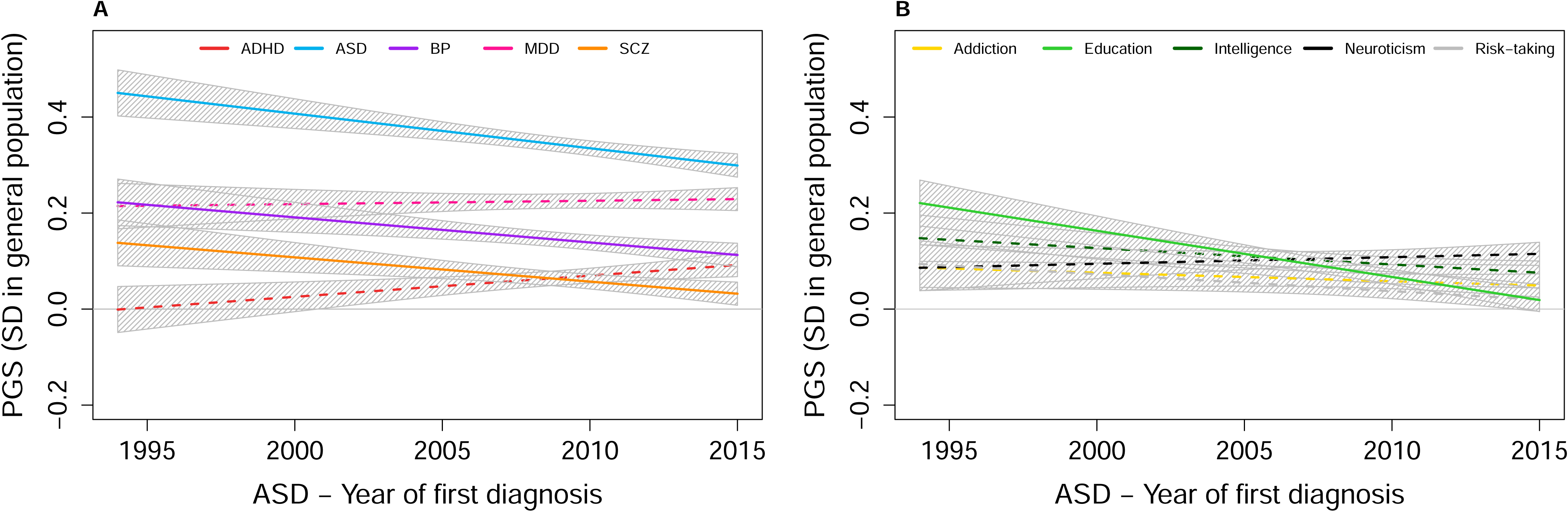
Genetic risk profile in ASD patients (N=17,071) reveals decreasing trends for year of diagnosis between 1994:2015 on x-axis. The y-axis shows polygenic score (PGS) level in standard deviations (SD) to the general population. Each line represents the regression results from each of the five psychiatric PGS in **A)** and five cognitive/behavioral PGS in **B)** predicted by year while adjusting for age and covariate, shown in **Table 1**. After Bonferroni correction (<0.05/20) solid lines are significant and dashed lines are not.

### Sensitivity Analysis

To test the robustness of our results we conducted several sensitivity analyses. For ADHD, adjustment of year by age was important, as the relationship appears related for MDD, Addiction and Neuroticism PGS (Supplementary Tables 4-13, Supplementary Figure 3). All results are robust to the choice of PGS training GWAS (Supplementary Table 24) and incorporating non-linear terms for year (Supplementary Table 25). The relationships between PGS and year were, in general, highly similar across sexes (Supplementary Tables 26). For ASD, adjustment of year by age had little impact (Supplementary Tables 14-23, Supplementary Figure 4) and results were similarly robust to training GWAS (Supplementary Table 27), non-linear trends (Supplementary Table 28), and sex-specific effects (Supplementary Tables 29). The results presented in Figure 3 and Figure 4 are highly stable.

## DISCUSSION

In this study, we show a decreasing impact of genetic risk on diagnoses of ASD and ADHD over a period in which the diagnostic rate has increased more than four fold. We interpret these results as consistent with prior studies suggesting a broadening of diagnostic criteria is the most plausible driver of increasing rates of both diagnoses.

Our approach is informative for differentiating among certain higher-level hypotheses, but we are limited in our ability to draw strong conclusions as to why a lowering threshold appears most consistent with our data. For example, the genetic risk profile can be reduced by mis-or over-diagnosis^13,15–17^. Here, individuals without ADHD or ASD (and thus low genetic risk) or milder presentations (assuming severity and genetic risk are correlated^42,43^) would lower the mean risk in genetic profiles of cases. An equivalent framing is that a lowering threshold is occurring because previously only the most severely affected individuals were diagnosed, as we note that although the profile has decreased, the PGS levels remain significantly higher than observed in the general population. Likewise, an increased burden of environmental causes could be consistent with a lowering threshold, as our study can only inform on the *genetic* threshold. For example, increased exposure to screens and social media^20^ or changes to early education environments^21^ have been proposed as risk factors, although evidence is neither consistently reported, nor required to explain the current trends. It is difficult to disambiguate among these hypotheses retrospectively, however, intervention studies, prospective longitudinal cohorts, and extracting detailed measures from, e.g., clinical notes could help.

Evolving nosology and changing clinical practice around adult diagnosis and recognition in females is reflected in time trends of the diagnosed population^3^, and as such we adjusted for age in our models and tested for sex interaction. While we did not observe sex differences in the genetic risk profile over time, the PGS associations for age revealed a transition with increasing age towards less neurodevelopmental and more internalizing and substance use genetic risk, but no changes in the index disorder PGS, consistent with our previous results for ADHD^32^. This preliminary finding suggests that the genetic relationships with other clinical features change with age, which could be explored in future studies.

This work significantly extends the limited body of work relating genetic risk to diagnostic time trends (i.e., year) in ADHD and ASD. Our results draw different conclusions than two previous studies using Swedish twin registers, suggesting genetic and environmental contributions to ADHD^29^ and ASD^28^ have not changed markedly over time. We believe direct measures of genetics, e.g., PGS, provide a more powerful index and this difference in design may be critical. Our present work adds significantly to our previous work^27^ in which we observed a decreased index PGS for ADHD and ASD diagnosed in individuals from more recent birth cohorts. We confirm initial support of lowering threshold with consistent evidence from multiple PGS and reveal unique associations to age.

Our results have important implications for the design, interpretation, and application of large-scale genetic epidemiological and gene-mapping studies. Few^28–30^ have considered the year of first diagnosis as a contributor to differences among cohorts. Our results here and previously reported changes^3^ over time in epidemiological or clinical characteristics, e.g., prevalence, age, sex ratio, etc., suggest time trends could contribute to cross-cohort heterogeneity. This may be especially true when considering age of onset or first diagnosis, as there are well-described time trends across all disorders^3^ and age (i.e., age at diagnosis) can be confounded with period (i.e., year of diagnosis) and cohort (i.e., birth year) in ways that are difficult to disambiguate^41^. Such differences could impact GWAS, which map different polygenes, if, for example, cases of ADHD or ASD were sampled using diagnoses made in different year windows. Speculatively, diagnostic time trends could also impact downstream applications of GWAS, e.g., PGS associations with other outcomes, GWAS revealing different patterns of genetic correlations, or different pathways being enriched for polygenic burden. As meta-studies become broader, more inclusive, and aggregate across longer time spans, understanding how time trends can impact on our inferences into etiology and predictive ability is important.

Our study has at least four limitations. First, the data from Danish registers do not include diagnosis made in primary care or by psychiatrists in specialized care outside hospital in-and out-patient facilities. Second, birth year truncation is embedded in the iPSYCH case-cohort design, which should be considered when generalizing findings. However, we note our previous work^30^ included restrictions to limit this and results are consistent. Third, the iPSYCH case-cohort is relatively young (i.e., few above age 35), which limits our ability to study a full life-course and earlier birth cohorts, although the recent introduction of ADHD and ASD and their typically early age at diagnosis are reassuring for our design. Fourth, our analysis was limited to Denmark using individuals of broadly northern European ancestry. Given the interest in this topic, replication to other populations, countries, societies, ethnic groups and healthcare settings is important.

## CONCLUSION

Our study offers a new perspective for on-going debates about increasing diagnostic rates of ASD and ADHD. By analyzing genetic risk profiles across two decades of diagnoses in a large population-based case-cohort, we demonstrate that individuals diagnosed more recently have systematically lower genetic risk. These findings suggest that clinical practice and diagnostic criteria have changed over time to include a broader phenotypic manifestation of ASD and ADHD.

## CONTRIBUTIONS

Conceptualization, S.L & A.J.S.; Methodology, S.L., M.L.L., M.D.K, O.P-R & A.J.S.; Formal Analysis, S.L., M.D.K., A.J.S.; Resources, iPSYCH Study Consortium, T.W.; Data Curation, S.L., O.S.H., C.A., B.J.V., M.L., J.R.G., H.O., iPSYCH Study Consortium, A.J.S.; Writing – Original Draft, S.L., A.J.S.; Writing – Review and Editing, S.L., M.L.L., M.D.K., K.-L.G.H, J.Ø.S, A.B., E.A., T.W., K.S.K., O.P-R, A.J.S.; Visualization, S.L., A.J.S.; Supervision, K.S.K., O.P-R., A.J.S.

## DATA SHARING

Access to individual-level Denmark data is governed by Danish authorities. These include the Danish Data Protection Agency, the Danish Health Data Authority, the Ethical Committee, and Statistics Denmark. Each scientific project must be approved before initiation, and approval is granted to a specific Danish research institution. Researchers at Danish research institutions may obtain the relevant approval and data. International researchers may gain data access if governed by a Danish research institution having needed approval and data access. R code for data analysis is available from corresponding author S.L. upon request. Simulation code is available online (https://github.com/sonjalabianca/Time-Trends).

## Supporting information

Supplementary Tabels

## Data Availability

Access to individual-level Denmark data is governed by Danish authorities. These include the Danish Data Protection Agency, the Danish Health Data Authority, the Ethical Committee, and Statistics Denmark. Each scientific project must be approved before initiation, and approval is granted to a specific Danish research institution. Researchers at Danish research institutions may obtain the relevant approval and data. International researchers may gain data access if governed by a Danish research institution having needed approval and data access.

## ACKNOWLEDGEMENTS

The project was specifically supported by the Research Fund of the Mental Health Services – Capital Region of Denmark R4A92 to S.L. M.D.K received funding from the Lundbeck Foundation (R450-2023-1447). O.P-R. received funding from Independent Research Fund Denmark (2066-00009B and 4309-00026B) AJS received funding from the Lundbeck Foundation (R335-2019-2318). A.J.S., and K.S.K. are supported by the National Institute of Mental Health R01MH130581. B.J.V. is supported by the Lundbeck Foundation (R335-2019-2339) and Independent Research Fund Denmark (2034-00241B). The iPSYCH team was supported by grants from the Lundbeck Foundation (R102-A9118, R155-2014-1724, and R248-2017-2003), NIH/NIMH (1U01MH109514-01 and 1R01MH124851-01) and the Universities and University Hospitals of Aarhus and Copenhagen. The Danish National Biobank resource was supported by the Novo Nordisk Foundation. High-performance computer capacity for handling and statistical analysis of iPSYCH data on the GenomeDK HPC facility was provided by the Center for Genomics and Personalized Medicine and the Centre for Integrative Sequencing, iSEQ, Aarhus University, Denmark. The funding sources played no part in the design and conduct of the study, analyses, interpretation of the data, or decision to submit the manuscript for publication. The authors thank Nicholas Schork for feedback and discussions.

