## Supplementary Note and Figures for "As rates of ASD and ADHD rise, genetic contributions fall: Evidence for widening diagnostic criteria"

### SUPPLEMENTARY NOTE & FIGURES

#### Supplementary Note

A population of 500,000 individuals was simulated in which two disorders (i.e., A and B) arise from a liability threshold model of disease(Falconer 1965). For both disorders the heritability ($h^2$=0.8) and variance explained by a hypothetical polygenic score (PGS, $R^2_{PGS}$=0.1) were fixed, but varying levels of true genetic correlation ($r_g$) between disorders A and B ($r_G$= 0, 0.25, 0.5, 0.75) was considered. For each genetic correlation scenario, the expected polygenic profiles (i.e., mean PGS for disorder A and disorder B) of individuals diagnosed with disorder A were simulated under three different generative models of potential causal mechanism : 1) lowering threshold, 2) shifting boundaries, and 3) better detection. In all three models the prevalence of disorder A increased (k=0.02, 0.025, 0.03, 0.035, 0.04). For the lowering threshold model a) the prevalence increased by lowering the liability threshold needed to receive a diagnosis. For the shifting boundaries model b), the prevalence increased by assigning disorder A to an increasing number of cases of disorder B (0.0, 0.1, 0.2, 0.3, 0.4, 0.5). For better detection c), the prevalence increased by lessening the number of true cases of disorder A that were identified as controls (0.5, 0.4, 0.3, 0.2, 0.1, 0.0). Simulation code is available online (https://github.com/sonjalabianca/Time-Trends).

a) Lowering threshold

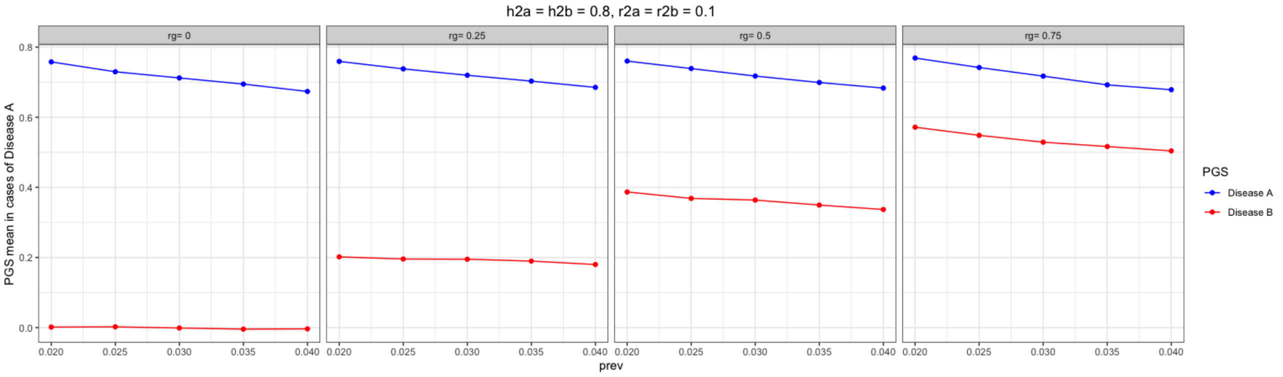


b) Shifting boundaries

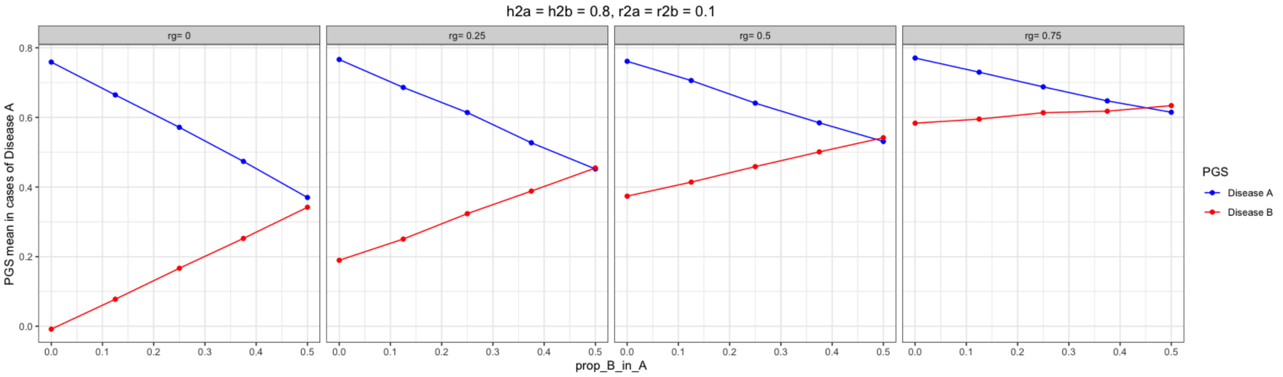

c) Better detection

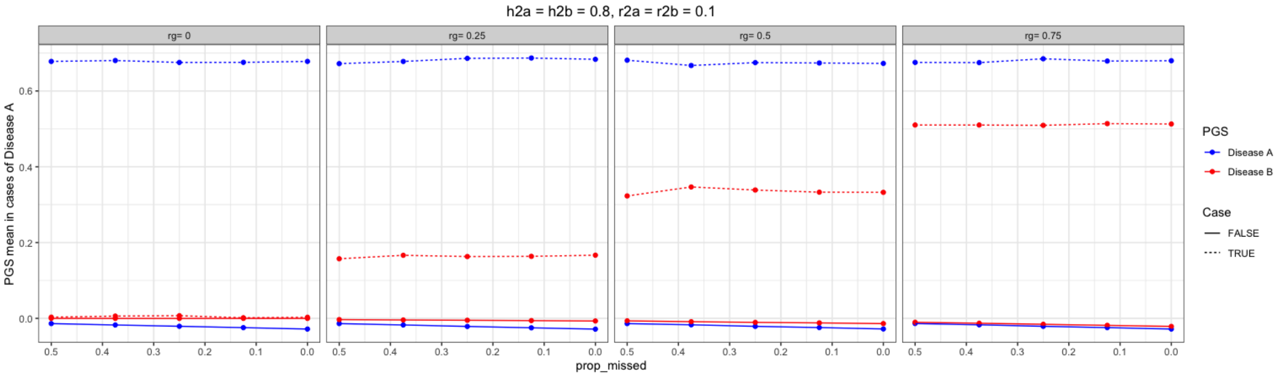

**Supplementary Figure 1.** Genetic risk profile in ADHD patients N=20,111 reveals diverse trends for age (*age at diagnosis*) between 1:35 on x-axis bottom. The y-axis shows polygenic score (PGS) levels in standard deviations (SD) to the general population. Each line represents the regression results from each of the five psychiatric PGS in **A)** and five cognitive/behavioral PGS in **B)** predicted by year while adjusting for age and covariate (Supplementary Tables 4-13). After Bonferroni correction (<0.05/20) solid lines are significant and dashed lines are not.

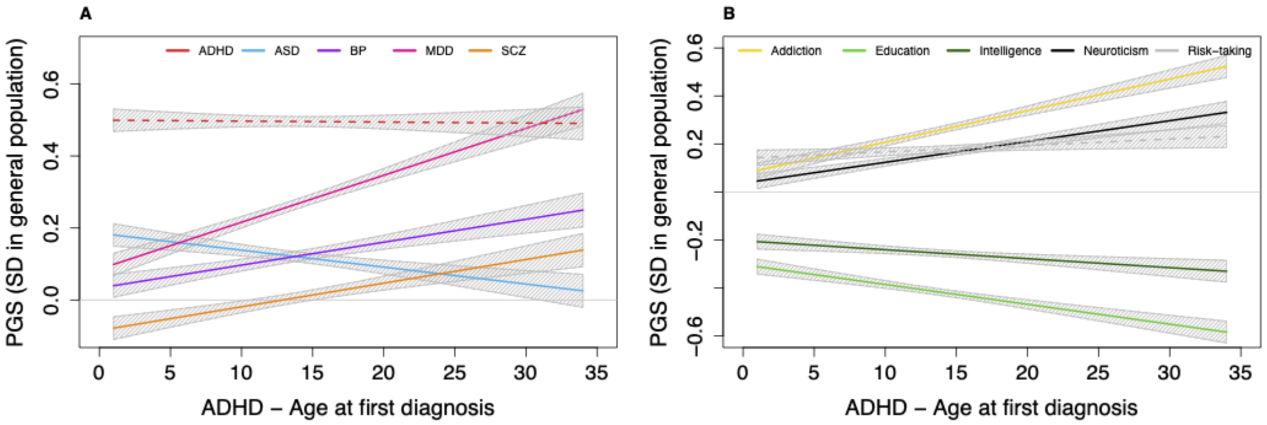

**Supplementary Figure 2.** Genetic risk profile in ASD patients N=17,071 reveals diverse trends for age (*age at diagnosis*) between 1:35 on x-axis bottom. The y-axis shows polygenic score level in standard deviations (SD) to the general population.Each line represents the regression results from each of the five psychiatric PGS in **A)** and five cognitive/behavioral PGS in **B)** predicted by year while adjusting for age and covariate (Supplementary Tables 17-26) **.** After Bonferroni correction (<0.05/20) solid lines are significant and dashed lines are not.

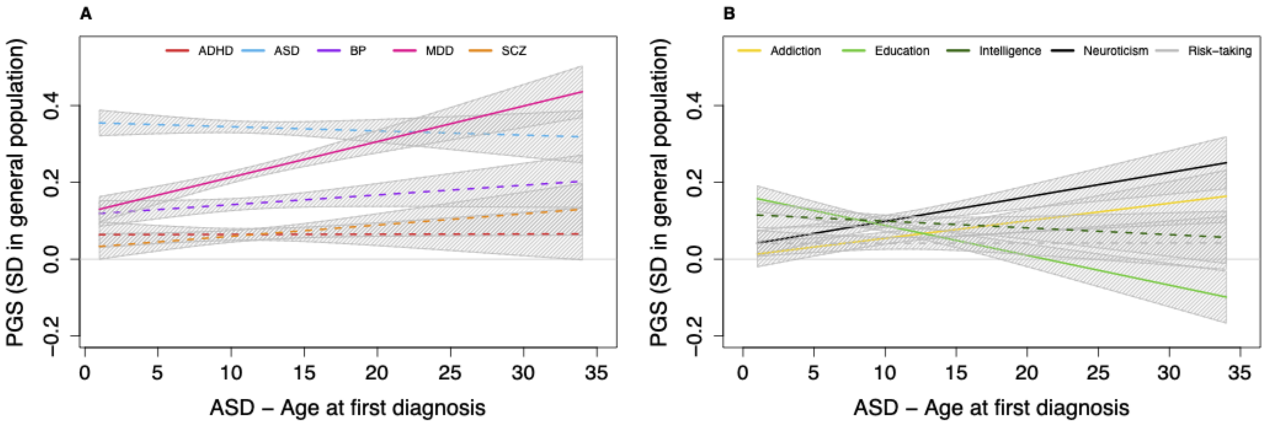

**Supplementary Figure 3.** Adjusting year by age can change the direction of effects in ADHD. y-axis=estimated betas for *year of diagnosis* in linear regression testing PGS~*year of diagnosis* + covariates (10 principal components + sex); x-axis=estimated betas for *year of diagnosis* in linear regression testing each PGS~*year of diagnosis* + *age of diagnosis* + covariates. Abbreviations: PGS=polygenic scores; ADHD=attention deficit hyperactivity disorder; ASD=autism spectrum disorder; BP=bipolar disorder; MDD=major depressive disorder; SCZ=schizophrenia; IQ=intelligence quotient

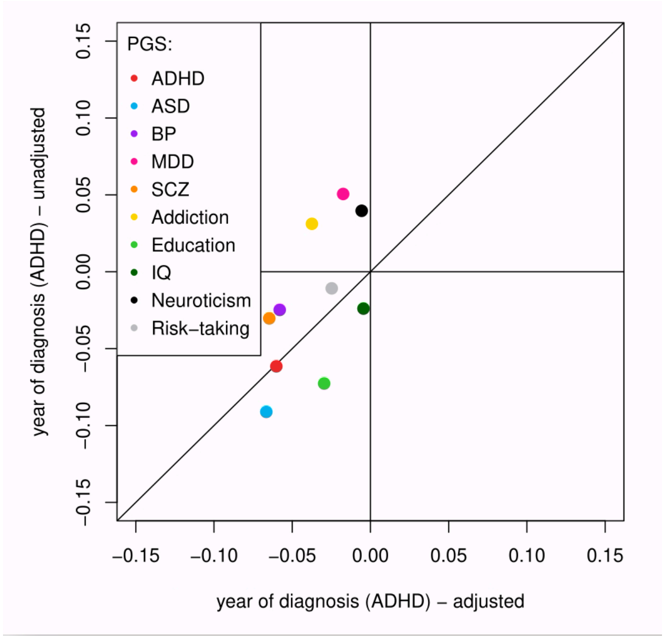

**Supplementary Figure 4.** Adjustment of year by age can change the magnitude of effects in ASD. y-axis=estimated betas for *year of diagnosis* in linear regression testing PGS~*year of diagnosis* + covariates (10 principal components + sex); x-axis=estimated betas for *year of diagnosis* in linear regression testing each PGS~*year of diagnosis* + *age of diagnosis* + covariates. Abbreviations: PGS=polygenic scores; ADHD=attention deficit hyperactivity disorder; ASD=autism spectrum disorder; BP=bipolar disorder; MDD=major depressive disorder; SCZ=schizophrenia; IQ=intelligence quotient.

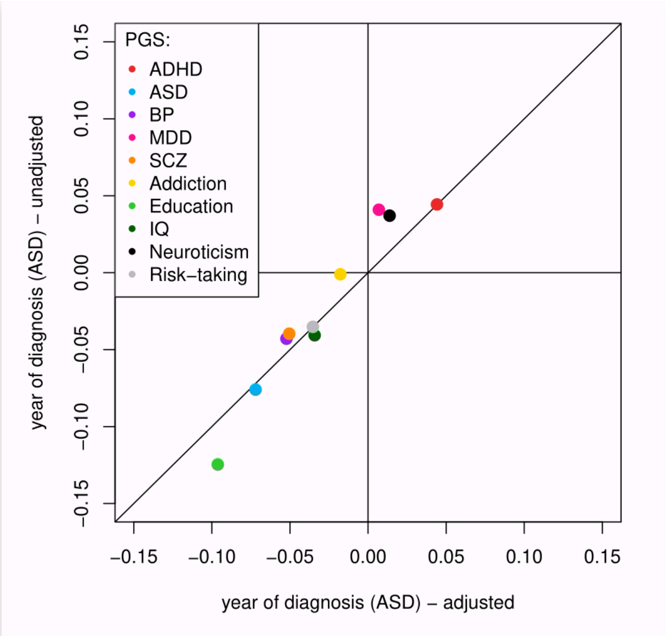
