## Supplementary Tabels for "As rates of ASD and ADHD rise, genetic contributions fall: Evidence for widening diagnostic criteria"

**Index of Supplementary Tables**

| | |
|---|---|
| ST1 | List of discovery GWAS for genetic risk profile |
| ST2 | Descriptiv statistics for ADHD patients in iPSYCH and the Danish Population |
| ST3 | Descriptiv statistics for ASD patients in iPSYCH and the Danish Population |
| ST4 | Year, age and cohort regression models unadjusted and adjusted for ADHD PGS in ADHD |
| ST5 | Year, age and cohort regression models unadjusted and adjusted for ASD PGS in ADHD |
| ST6 | Year, age and cohort regression models unadjusted and adjusted for BP PGS in ADHD |
| ST7 | Year, age and cohort regression models unadjusted and adjusted for MDD PGS in ADHD |
| ST8 | Year, age and cohort regression models unadjusted and adjusted for SCZ PGS in ADHD |
| ST9 | Year, age and cohort regression models unadjusted and adjusted for Addiction PGS in ADHD |
| ST10 | Year, age and cohort regression models unadjusted and adjusted for Education PGS in ADHD |
| ST11 | Year, age and cohort regression models unadjusted and adjusted for Intelligence PGS in ADHD |
| ST12 | Year, age and cohort regression models unadjusted and adjusted for Risk-taking PGS in ADHD |
| ST13 | Year, age and cohort regression models unadjusted and adjusted for Neuroticism PGS in ADHD |
| ST14 | Year, age and cohort regression models unadjusted and adjusted for ASD PGS in ASD |
| ST15 | Year, age and cohort regression models unadjusted and adjusted for ADHD PGS in ASD |
| ST16 | Year, age and cohort regression models unadjusted and adjusted for BP PGS in ASD |
| ST17 | Year, age and cohort regression models unadjusted and adjusted for MDD PGS in ASD |
| ST18 | Year, age and cohort regression models unadjusted and adjusted for SCZ PGS in ASD |
| ST19 | Year, age and cohort regression models unadjusted and adjusted for Addiction PGS in ASD |
| ST20 | Year, age and cohort regression models unadjusted and adjusted for Education PGS in ASD |
| ST21 | Year, age and cohort regression models unadjusted and adjusted for Intelligence PGS in ASD |
| ST22 | Year, age and cohort regression models unadjusted and adjusted for Risk-taking PGS in ASD |
| ST23 | Year, age and cohort regression models unadjusted and adjusted for Neuroticism PGS in ASD |
| ST24 | Year and age mutuy adjusted models for five psychiatric PGS excl. iPSYCH in ADHD |
| ST25 | Comparing linear and nonlinar effects of year and age in ADHD |
| ST26 | Sex interaction with year in mutuy adjusted models for 10 PGS in ADHD |
| ST27 | Year and age mutuy adjusted models for five psychiatric PGS excl. iPSYCH in ASD |
| ST28 | Sex interaction with year in mutuy adjusted models for 10 PGS in ASD |
| ST29 | Comparing linear and nonlinar effects of year and age in ASD |
| ST30 | iPSYCH Study Consortium banner authors |

**Supplementary Table 1. List of discovery GWAS for polygenic profile.** Abbreviations: GWAS= genome wide association study; ADHD=attention deficit hyperactivity disorder; ASD=autism spectrum disorder; BP=bipolar disorder; MDD=major depressiv disorder; SCZ=schizophrenia; EA=educational attainment; IQ=intelligent quotient.

| Disorder/Trait | Reference of Discovery GWAS |
|---|---|
| ADHD | Demontis, D. *et al.* Discovery of the first genome-wide significant risk loci for attention deficit/hyperactivity disorder. *Nat. Genet.* (2018) doi:10.1038/s41588-018-0269-7. |
| ASD | Grove, J. *et al.* Identification of common genetic risk variants for autism spectrum disorder. *Nat. Genet.* **51**, 431–444 (2019). |
| BP | Stahl, E. A. *et al.* Genome-wide association study identifies 30 loci associated with bipolar disorder. *Nature genetics* **51**, (2019). |
| MDD | Howard, D. M. *et al.* Genome-wide meta-analysis of depression identifies 102 independent variants and highlights the importance of the prefrontal brain regions. *Nat. Neurosci.* **22**, 343–352 (2019). |
| SCZ | Ripke, S. *et al.* Biological insights from 108 schizophrenia-associated genetic loci. *Nature* **511**, 421–427 (2014). |
| EA | Okbay, A. *et al.* Polygenic prediction of educational attainment within and between families from genome-wide association analyses in 3 million individuals. *Nat. Genet.* **54**, 437–449 (2022). |
| IQ | Savage, J. E. *et al.* Genome-wide association meta-analysis in 269,867 individuals identifies new genetic and functional links to intelligence. *Nat. Genet.* **50**, 912–919 (2018). |
| Risk-taking | Karlsson Linnér, R. *et al.* Genome-wide association analyses of risk tolerance and risky behaviors in over 1 million individuals identify hundreds of loci and shared genetic influences. *Nat. Genet.* **51**, 245–257 (2019). |
| Neuroticism | Nagel, M. *et al.* Meta-analysis of genome-wide association studies for neuroticism in 449,484 individuals identifies novel genetic loci and pathways. (2018) doi:10.1038/s41588-018-0151-7. |
| Addiction | Hatoum, A. S. *et al.* The addiction risk factor: A unitary genetic vulnerability characterizes substance use disorders and their associations with common correlates. *Neuropsychopharmacology* **47**, 1739–1745 (2022). |

**Supplementary Table 2. Descriptive statistics for ADHD patients (ICD10: F90.0) in iPSYCH N=20,111 and the Danish poulation N=34,516.** Period=Year of first diagnosis; N=Number of patients; POP=Danish population; Age=Age at first diganosis; SD=Standard deviations.; NA=Not available due to data protection regulations restricting information to cells >=5 indidivuals.

| Period | N_iPSYCH | N_POP | Age_iPSYCH (mean) | Age_iPSYCH (SD) | Age_POP (mean) | Age_POP (SD) | Females_iPSYCH (%) | Females_POP (%) |
|---|---|---|---|---|---|---|---|---|
| 1994 | 35 | 100 | 7.54 | 2.90 | 9.19 | 8.34 | NA | 14 |
| 1995 | 111 | 255 | 8.23 | 2.52 | 8.75 | 3.09 | 13.51 | 16.47 |
| 1996 | 87 | 213 | 8.13 | 2.71 | 8.88 | 6.34 | 11.49 | 18.31 |
| 1997 | 109 | 193 | 7.93 | 2.54 | 8.68 | 3.97 | 11.93 | 15.54 |
| 1998 | 112 | 196 | 7.98 | 2.78 | 9.35 | 6.45 | 12.50 | 11.22 |
| 1999 | 161 | 263 | 8.48 | 2.93 | 9.26 | 6.13 | 9.94 | 8.75 |
| 2000 | 162 | 288 | 8.23 | 3.11 | 8.76 | 3.54 | 20.99 | 17.71 |
| 2001 | 231 | 328 | 8.55 | 3.37 | 8.88 | 5.34 | 15.15 | 18.6 |
| 2002 | 263 | 407 | 8.66 | 3.63 | 9.12 | 5.54 | 12.93 | 14.5 |
| 2003 | 366 | 566 | 9.03 | 3.79 | 9.55 | 4.42 | 15.85 | 15.72 |
| 2004 | 494 | 787 | 9.62 | 3.93 | 11.09 | 7.54 | 16.19 | 16.14 |
| 2005 | 582 | 894 | 10.39 | 4.13 | 11.65 | 6.76 | 20.62 | 21.14 |
| 2006 | 740 | 1228 | 10.52 | 4.77 | 12.66 | 8.52 | 19.05 | 22.88 |
| 2007 | 902 | 1542 | 11.53 | 5.41 | 14.39 | 9.66 | 25.50 | 26.13 |
| 2008 | 1412 | 2417 | 12.41 | 5.77 | 15.62 | 10.14 | 26.13 | 28.67 |
| 2009 | 1568 | 2969 | 13.60 | 6.17 | 16.56 | 10.53 | 28.76 | 28.39 |
| 2010 | 1891 | 3302 | 14.23 | 6.42 | 18.2 | 11.16 | 29.88 | 29.16 |
| 2011 | 1954 | 3361 | 14.71 | 6.82 | 19.19 | 11.45 | 30.76 | 31.54 |
| 2012 | 2241 | 3706 | 16.02 | 7.06 | 20.31 | 11.54 | 33.73 | 35.02 |
| 2013 | 2267 | 3843 | 15.56 | 7.11 | 19.84 | 11.42 | 35.55 | 35.68 |
| 2014 | 2261 | 3896 | 16.25 | 7.34 | 21.01 | 12.02 | 36.67 | 36.58 |
| 2015 | 2162 | 3762 | 15.92 | 7.40 | 19.5 | 11.98 | 36.17 | 36.28 |

**Supplementary Table 3. Descriptive statistics for ASD patients (ICD10: F85.0;F84.1;F84.5;F84.8;F84.9) in iPSYCH N=17,071 and the entire Danish poulation N=26,326.** Period=Year of first diagnosis; N=Number of patients; POP=Danish population; Age=Age at first diganosis; SD=Standard deviations.

| Period | N_iPSYCH | N_POP | Age_iPSYCH (mean) | Age_iPSYCH (SD) | Age_POP (mean) | Age_POP (SD) | Females_iPSYCH (%) | Females_POP (%) |
|---|---|---|---|---|---|---|---|---|
| 1994 | 74 | 198 | 6.73 | 3.22 | 14.57 | 12.13 | 14.86 | 18.69 |
| 1995 | 187 | 367 | 7.42 | 3.30 | 11.65 | 9.59 | 16.04 | 21.53 |
| 1996 | 191 | 366 | 7.95 | 3.26 | 12.01 | 8.80 | 17.80 | 19.4 |
| 1997 | 233 | 405 | 7.89 | 3.62 | 11.23 | 9.37 | 14.59 | 18.52 |
| 1998 | 270 | 438 | 7.74 | 3.82 | 9.67 | 7.49 | 12.22 | 16.21 |
| 1999 | 328 | 591 | 8.02 | 3.91 | 10.47 | 8.06 | 16.16 | 18.61 |
| 2000 | 384 | 644 | 7.52 | 3.89 | 9.51 | 7.50 | 15.36 | 18.32 |
| 2001 | 407 | 699 | 8.22 | 4.04 | 10.79 | 9.22 | 13.76 | 16.17 |
| 2002 | 455 | 742 | 8.56 | 4.27 | 10.27 | 8.47 | 15.60 | 16.58 |
| 2003 | 440 | 714 | 8.84 | 4.44 | 10.8 | 9.60 | 15.68 | 17.23 |
| 2004 | 556 | 865 | 9.26 | 4.58 | 11.75 | 10.03 | 18.88 | 19.77 |
| 2005 | 589 | 925 | 9.19 | 4.76 | 10.91 | 8.30 | 20.20 | 19.89 |
| 2006 | 754 | 1107 | 9.73 | 4.79 | 10.92 | 7.36 | 21.88 | 21.05 |
| 2007 | 795 | 1200 | 9.92 | 4.87 | 11.98 | 8.92 | 20.88 | 21.33 |
| 2008 | 1064 | 1566 | 10.35 | 4.98 | 11.66 | 8.00 | 21.80 | 21.2 |
| 2009 | 1065 | 1666 | 11.09 | 5.13 | 12.58 | 9.12 | 24.88 | 21.91 |
| 2010 | 1224 | 1806 | 11.08 | 5.48 | 12.09 | 8.11 | 24.18 | 22.7 |
| 2011 | 1355 | 1909 | 11.61 | 5.55 | 12.87 | 8.88 | 24.58 | 24.99 |
| 2012 | 1404 | 1984 | 12.31 | 5.89 | 13.63 | 9.78 | 28.13 | 26.81 |
| 2013 | 1652 | 2415 | 12.67 | 5.63 | 13.51 | 8.83 | 29.84 | 28.9 |
| 2014 | 1796 | 2738 | 13.73 | 5.81 | 14.32 | 9.43 | 31.46 | 29.62 |
| 2015 | 1848 | 2981 | 14.46 | 5.75 | 14.59 | 9.95 | 33.12 | 30.43 |

**Supplementary Table 4. Linear regression models for ADHD polygenic score in ADHD patients N=20,111**. Abbreviations: PC=principal component; Year of diagnosis (ADHD)=first recorded ADHD diagnosis for each individuals between 1994:2016 obtained from Danish national registers; Age at diagnosis (ADHD)=age at first recorded ADHD diagnosis for each individual between 1:35 obtained from Danish national health registers; Year of birth= between 1981:2008 obtained from Danish national health registers.

| Predictor | Model 0 | Model 1 | Model 2 | Model 3 | Model 4 | Model 5 |
|---|---|---|---|---|---|---|
| **(Intercept)** | 0.52 (0.49:0.54), <0.001 | 0.62 (0.56:0.68), <0.001 | 0.62 (0.56:0.69), <0.001 | 0.54 (0.50:0.58), <0.001 | 0.71 (0.60:0.81), <0.001 | 0.53 (0.49:0.57), <0.001 |
| **PC1** | 8.08 (3.45:12.70), 0.001 | 8.03 (3.41:12.66), 0.001 | 8.01 (3.38:12.64), 0.001 | 7.93 (3.29:12.56), 0.001 | 8.01 (3.37:12.64), 0.001 | 8.17 (3.53:12.80), 0.001 |
| **PC2** | 1.93 (-2.53:6.39), 0.396 | 2.00 (-2.46:6.45), 0.379 | 1.99 (-2.46:6.45), 0.380 | 1.92 (-2.54:6.37), 0.399 | 2.00 (-2.45:6.46), 0.378 | 1.95 (-2.51:6.41), 0.391 |
| **PC3** | -7.70 (-12.34:-3.05), 0.001 | -7.86 (-12.50:-3.21), 0.001 | -7.86 (-12.50:-3.21), 0.001 | -7.71 (-12.36:-3.06), 0.001 | -7.85 (-12.50:-3.21), 0.001 | -7.71 (-12.36:-3.07), 0.001 |
| **PC4** | 3.49 (-0.99:7.97), 0.126 | 3.47 (-1.00:7.95), 0.128 | 3.48 (-1.00:7.96), 0.128 | 3.52 (-0.96:8.00), 0.123 | 3.47 (-1.00:7.95), 0.128 | 3.47 (-1.01:7.94), 0.129 |
| **PC5** | 1.38 (-3.10:5.86), 0.546 | 1.46 (-3.02:5.94), 0.522 | 1.46 (-3.02:5.94), 0.522 | 1.41 (-3.07:5.89), 0.538 | 1.46 (-3.02:5.94), 0.522 | 1.38 (-3.10:5.86), 0.547 |
| **PC6** | -2.64 (-7.10:1.82), 0.245 | -2.54 (-7.00:1.92), 0.264 | -2.54 (-7.00:1.92), 0.264 | -2.60 (-7.06:1.86), 0.252 | -2.54 (-7.00:1.92), 0.264 | -2.65 (-7.11:1.80), 0.243 |
| **PC7** | 5.98 (1.51:10.44), 0.009 | 6.01 (1.55:10.47), 0.008 | 6.01 (1.55:10.48), 0.008 | 5.99 (1.53:10.46), 0.009 | 6.02 (1.56:10.48), 0.008 | 5.97 (1.51:10.44), 0.009 |
| **PC8** | -4.36 (-8.82:0.10), 0.055 | -4.35 (-8.80:0.11), 0.056 | -4.35 (-8.80:0.11), 0.056 | -4.36 (-8.82:0.10), 0.055 | -4.35 (-8.81:0.10), 0.056 | -4.36 (-8.82:0.10), 0.055 |
| **PC9** | 19.47 (14.97:23.97), <0.001 | 19.44 (14.94:23.94), <0.001 | 19.44 (14.94:23.93), <0.001 | 19.44 (14.94:23.94), <0.001 | 19.44 (14.94:23.93), <0.001 | 19.48 (14.98:23.98), <0.001 |
| **PC10** | 5.03 (0.53:9.52), 0.028 | 5.01 (0.52:9.51), 0.029 | 5.01 (0.52:9.51), 0.029 | 5.03 (0.53:9.52), 0.028 | 5.02 (0.53:9.51), 0.029 | 5.03 (0.53:9.52), 0.028 |
| **Sex** | -0.03 (-0.06:-0.00), 0.030 | -0.04 (-0.07:-0.01), 0.007 | -0.04 (-0.07:-0.01), 0.007 | -0.04 (-0.07:-0.01), 0.016 | -0.04 (-0.07:-0.01), 0.007 | -0.03 (-0.06:-0.00), 0.040 |
| **Year of diagnosis (ADHD)** | | -0.06 (-0.09:-0.03), <0.001 | -0.06 (-0.09:-0.03), 0.001 | | | |
| **Age at diagnosis (ADHD)** | | | -0.00 (-0.02:0.02), 0.811 | -0.01 (-0.04:0.01), 0.156 | -0.06 (-0.10:-0.03), <0.001 | |
| **Year of birth** | | | | | -0.06 (-0.10:-0.03), 0.001 | -0.01 (-0.03:0.01), 0.351 |

**Supplementary Table 5. Linear regression models for ASD polygenic score in ADHD patients N=20,111**. Abbreviations: PC=principal component; Year of diagnosis (ADHD)=first recorded ADHD diagnosis for each individuals between 1994:2016 obtained from Danish national registers; Age at diagnosis (ADHD)=age at first recorded ADHD diagnosis for each individual between 1:35 obtained from Danish national health registers; Year of birth= between 1981:2008 obtained from Danish national health registers.

| **Predictors** | **Model 0** | **Model 1** | **Model 2** | **Model 3** | **Model 4** | **Model 5** |
|---|---|---|---|---|---|---|
| **(Intercept)** | 0.14 (0.11:0.16), p<0.001 | 0.29 (0.23:0.35), p<0.001 | 0.33 (0.27:0.40), p<0.001 | 0.24 (0.20:0.28), p<0.001 | 0.41 (0.30:0.51), p<0.001 | 0.10 (0.06:0.14), p<0.001 |
| **PC1** | 12.00 (7.35:16.65), p<0.001 | 11.94 (7.29:16.58), p<0.001 | 11.48 (6.83:16.13), p<0.001 | 11.39 (6.74:16.04), p<0.001 | 11.47 (6.82:16.12), p<0.001 | 11.74 (7.09:16.40), p<0.001 |
| **PC2** | 9.76 (5.28:14.24), p<0.001 | 9.86 (5.39:14.34), p<0.001 | 9.80 (5.32:14.27), p<0.001 | 9.71 (5.24:14.18), p<0.001 | 9.80 (5.33:14.27), p<0.001 | 9.70 (5.23:14.18), p<0.001 |
| **PC3** | -6.25 (-10.92:-1.58), p=0.009 | -6.49 (-11.15:-1.82), p=0.006 | -6.46 (-11.13:-1.80), p=0.007 | -6.30 (-10.97:-1.64), p=0.008 | -6.45 (-11.11:-1.78), p=0.007 | -6.20 (-10.87:-1.53), p=0.009 |
| **PC4** | -0.11 (-4.61:4.39), p=0.962 | -0.13 (-4.63:4.36), p=0.954 | -0.03 (-4.52:4.46), p=0.989 | 0.01 (-4.48:4.51), p=0.996 | -0.03 (-4.53:4.46), p=0.988 | -0.04 (-4.54:4.45), p=0.985 |
| **PC5** | -4.62 (-9.12:-0.12), p=0.044 | -4.50 (-9.00:-0.00), p=0.050 | -4.45 (-8.95:0.05), p=0.052 | -4.51 (-9.01:-0.02), p=0.049 | -4.46 (-8.95:-0.04), p=0.052 | -4.61 (-9.11:-0.11), p=0.043 |
| **PC6** | 4.04 (-0.44:8.51), p=0.077 | 4.18 (-0.29:8.66), p=0.067 | 4.27 (-0.20:8.75), p=0.061 | 4.20 (-0.28:8.68), p=0.066 | 4.26 (-0.21:8.74), p=0.062 | 4.07 (-0.41:8.54), p=0.075 |
| **PC7** | -16.99 (-21.47:-12.50), p<0.001 | -16.94 (-21.42:-12.46), p<0.001 | -16.90 (-21.38:-12.42), p<0.001 | -16.92 (-21.41:-12.44), p<0.001 | -16.89 (-21.38:-12.41), p<0.001 | -16.98 (-21.46:-12.49), p<0.001 |
| **PC8** | -3.54 (-8.02:0.94), p=0.121 | -3.52 (-8.00:0.96), p=0.123 | -3.54 (-8.01:0.94), p=0.121 | -3.55 (-8.03:0.92), p=0.120 | -3.54 (-8.02:0.93), p=0.120 | -3.55 (-8.03:0.93), p=0.120 |
| **PC9** | 7.66 (3.14:12.18), p=0.001 | 7.62 (3.10:12.13), p=0.001 | 7.56 (3.05:12.07), p=0.001 | 7.57 (3.05:12.09), p=0.001 | 7.56 (3.05:12.08), p=0.001 | 7.63 (3.11:12.15), p=0.001 |
| **PC10** | 4.84 (0.32:9.36), p=0.036 | 4.82 (0.31:9.33), p=0.036 | 4.82 (0.31:9.33), p=0.036 | 4.83 (0.32:9.35), p=0.036 | 4.83 (0.32:9.34), p=0.036 | 4.84 (0.33:9.35), p=0.036 |
| **Sex** | -0.03 (-0.06:0.00), p=0.080 | -0.04 (-0.07:-0.01), p=0.011 | -0.05 (-0.08:-0.02), p=0.001 | -0.05 (-0.08:-0.01), p=0.004 | -0.05 (-0.08:-0.02), p=0.002 | -0.03 (-0.06:-0.00), p=0.042 |
| **Year of diagnosis (ADHD)** | | -0.09 (-0.12:-0.06), p<0.001 | -0.07 (-0.10:-0.03), p<0.001 | | | |
| **Age at diagnosis (ADHD)** | | | -0.05 (-0.07:-0.03), p<0.001 | -0.06 (-0.08:-0.04), p<0.001 | -0.11 (-0.14:-0.07), p<0.001 | |
| **Year of birth** | | | | | -0.06 (-0.10:-0.03), p=0.001 | 0.03 (0.01:0.05), p=0.010 |

**Supplementary Table 6. Linear regression models for Bipolar (BP) polygenic score in ADHD patients N=20,111**. Abbreviations: PC=principal component; Year of diagnosis (ADHD)=first recorded ADHD diagnosis for each individuals between 1994:2016 obtained from Danish national registers; Age at diagnosis (ADHD)=age at first recorded ADHD diagnosis for each individual between 1:35 obtained from Danish national health registers; Year of birth= between 1981:2008 obtained from Danish national health registers.

| Predictor | Model 0 | Model 1 | Model 2 | Model 3 | Model 4 | Model 5 |
|---|---|---|---|---|---|---|
| **(Intercept)** | 0.15 (0.13:0.18), <0.001 | 0.20 (0.13:0.26), <0.001 | 0.14 (0.08:0.21), <0.001 | 0.07 (0.02:0.11), 0.004 | 0.22 (0.11:0.32), <0.001 | 0.24 (0.20:0.28), <0.001 |
| **PC1** | 11.22 (6.44:16.00), <0.001 | 11.21 (6.42:15.99), <0.001 | 11.82 (7.04:16.60), <0.001 | 11.74 (6.96:16.53), <0.001 | 11.81 (7.03:16.60), <0.001 | 11.79 (7.01:16.58), <0.001 |
| **PC2** | -1.93 (-6.53:2.67), 0.411 | -1.90 (-6.51:2.70), 0.418 | -1.81 (-6.41:2.79), 0.440 | -1.89 (-6.49:2.71), 0.421 | -1.81 (-6.41:2.79), 0.440 | -1.80 (-6.40:2.80), 0.442 |
| **PC3** | -1.62 (-6.42:3.18), 0.507 | -1.69 (-6.49:3.11), 0.491 | -1.72 (-6.51:3.08), 0.483 | -1.58 (-6.37:3.22), 0.519 | -1.71 (-6.50:3.09), 0.485 | -1.73 (-6.52:3.07), 0.481 |
| **PC4** | 3.56 (-1.06:8.19), 0.131 | 3.56 (-1.07:8.18), 0.132 | 3.42 (-1.20:8.04), 0.147 | 3.46 (-1.16:8.08), 0.142 | 3.42 (-1.20:8.04), 0.147 | 3.42 (-1.20:8.04), 0.147 |
| **PC5** | 0.91 (-3.72:5.54), 0.700 | 0.94 (-3.69:5.57), 0.690 | 0.88 (-3.75:5.50), 0.710 | 0.82 (-3.80:5.45), 0.728 | 0.87 (-3.75:5.50), 0.711 | 0.88 (-3.74:5.51), 0.708 |
| **PC6** | 1.48 (-3.13:6.08), 0.530 | 1.52 (-3.09:6.12), 0.518 | 1.40 (-3.20:6.00), 0.551 | 1.34 (-3.27:5.94), 0.569 | 1.39 (-3.21:6.00), 0.553 | 1.41 (-3.19:6.01), 0.549 |
| **PC7** | -7.10 (-11.71:-2.49), 0.003 | -7.08 (-11.69:-2.47), 0.003 | -7.13 (-11.74:-2.52), 0.002 | -7.15 (-11.76:-2.54), 0.002 | -7.13 (-11.73:-2.52), 0.002 | -7.12 (-11.73:-2.51), 0.002 |
| **PC8** | -4.17 (-8.77:0.44), 0.076 | -4.16 (-8.76:0.44), 0.077 | -4.14 (-8.74:0.46), 0.078 | -4.16 (-8.76:0.45), 0.077 | -4.15 (-8.75:0.45), 0.077 | -4.15 (-8.75:0.46), 0.077 |
| **PC9** | -4.73 (-9.38:-0.09), 0.046 | -4.74 (-9.39:-0.10), 0.045 | -4.67 (-9.31:-0.02), 0.049 | -4.66 (-9.30:-0.01), 0.049 | -4.67 (-9.31:-0.02), 0.049 | -4.67 (-9.31:-0.03), 0.049 |
| **PC10** | 2.36 (-2.28:7.00), 0.319 | 2.35 (-2.29:6.99), 0.321 | 2.35 (-2.29:6.99), 0.321 | 2.36 (-2.28:7.00), 0.318 | 2.36 (-2.28:7.00), 0.319 | 2.36 (-2.28:7.00), 0.319 |
| **Sex** | -0.05 (-0.08:-0.01), 0.004 | -0.05 (-0.08:-0.02), 0.002 | -0.03 (-0.07:-0.00), 0.032 | -0.03 (-0.06:0.00), 0.060 | -0.03 (-0.07:-0.00), 0.033 | -0.04 (-0.07:-0.00), 0.024 |
| **Year of diagnosis (ADHD)** | | -0.02 (-0.06:0.01), 0.152 | -0.06 (-0.09:-0.02), 0.001 | | | |
| **Age at diagnosis (ADHD)** | | | 0.06 (0.04:0.09), <0.001 | 0.05 (0.03:0.07), <0.001 | 0.01 (-0.03:0.04), 0.665 | |
| **Year of birth** | | | | | -0.06 (-0.09:-0.02), 0.002 | -0.06 (-0.08:-0.04), <0.001 |

**Supplementary Table 7. Linear regression models for Major depressive disorder (MDD) polygenic score in ADHD patients N=20,111**. Abbreviations: PC=principal component; Year of diagnosis (ADHD)=first recorded ADHD diagnosis for each individuals between 1994:2016 obtained from Danish national registers; Age at diagnosis (ADHD)=age at first recorded ADHD diagnosis for each individual between 1:35 obtained from Danish national health registers; Year of birth= between 1981:2008 obtained from Danish national health registers.

| Predictor | Model 0 | Model 1 | Model 2 | Model 3 | Model 4 | Model 5 |
|---|---|---|---|---|---|---|
| **(Intercept)** | 0.33 (0.31:0.36), <0.001 | 0.25 (0.18:0.31), <0.001 | 0.14 (0.08:0.20), <0.001 | 0.12 (0.07:0.16), <0.001 | 0.17 (0.06:0.27), 0.001 | 0.48 (0.44:0.52), <0.001 |
| **PC1** | 11.44 (6.82:16.06), <0.001 | 11.47 (6.85:16.09), <0.001 | 12.73 (8.13:17.34), <0.001 | 12.71 (8.10:17.32), <0.001 | 12.73 (8.13:17.34), <0.001 | 12.45 (7.84:17.06), <0.001 |
| **PC2** | -2.38 (-6.83:2.07), 0.294 | -2.44 (-6.88:2.01), 0.283 | -2.25 (-6.68:2.18), 0.319 | -2.27 (-6.71:2.16), 0.314 | -2.25 (-6.68:2.18), 0.320 | -2.15 (-6.59:2.28), 0.341 |
| **PC3** | -3.22 (-7.86:1.41), 0.173 | -3.09 (-7.73:1.55), 0.191 | -3.15 (-7.77:1.47), 0.181 | -3.11 (-7.73:1.51), 0.187 | -3.15 (-7.78:1.47), 0.181 | -3.41 (-8.03:1.22), 0.149 |
| **PC4** | 2.60 (-1.86:7.07), 0.253 | 2.62 (-1.85:7.09), 0.251 | 2.34 (-2.11:6.79), 0.303 | 2.35 (-2.10:6.80), 0.300 | 2.34 (-2.11:6.79), 0.303 | 2.35 (-2.11:6.80), 0.302 |
| **PC5** | 5.67 (1.20:10.15), 0.013 | 5.61 (1.14:10.08), 0.014 | 5.48 (1.02:9.93), 0.016 | 5.46 (1.01:9.92), 0.016 | 5.48 (1.02:9.93), 0.016 | 5.63 (1.17:10.09), 0.013 |
| **PC6** | -2.59 (-7.04:1.86), 0.254 | -2.67 (-7.12:1.78), 0.239 | -2.92 (-7.35:1.52), 0.197 | -2.93 (-7.37:1.50), 0.195 | -2.92 (-7.35:1.52), 0.197 | -2.71 (-7.15:1.72), 0.231 |
| **PC7** | -1.72 (-6.17:2.74), 0.450 | -1.75 (-6.20:2.71), 0.443 | -1.85 (-6.28:2.59), 0.415 | -1.85 (-6.29:2.59), 0.414 | -1.84 (-6.28:2.60), 0.416 | -1.76 (-6.20:2.69), 0.438 |
| **PC8** | -7.45 (-11.90:-3.00), 0.001 | -7.46 (-11.91:-3.01), 0.001 | -7.42 (-11.85:-2.99), 0.001 | -7.42 (-11.86:-2.99), 0.001 | -7.42 (-11.85:-2.99), 0.001 | -7.41 (-11.85:-2.98), 0.001 |
| **PC9** | 2.63 (-1.86:7.12), 0.251 | 2.65 (-1.84:7.14), 0.247 | 2.81 (-1.66:7.29), 0.218 | 2.82 (-1.66:7.29), 0.217 | 2.81 (-1.66:7.29), 0.218 | 2.74 (-1.74:7.22), 0.230 |
| **PC10** | 3.33 (-1.16:7.81), 0.146 | 3.34 (-1.14:7.82), 0.144 | 3.33 (-1.13:7.80), 0.144 | 3.34 (-1.13:7.81), 0.143 | 3.34 (-1.13:7.81), 0.143 | 3.33 (-1.15:7.80), 0.145 |
| **Sex** | -0.09 (-0.12:-0.06), <0.001 | -0.09 (-0.12:-0.06), <0.001 | -0.06 (-0.09:-0.03), <0.001 | -0.06 (-0.09:-0.03), <0.001 | -0.06 (-0.09:-0.03), <0.001 | -0.08 (-0.11:-0.05), <0.001 |
| **Year of diagnosis (ADHD)** | | 0.05 (0.02:0.08), 0.002 | -0.02 (-0.05:0.02), 0.321 | | | |
| **Age at diagnosis (ADHD)** | | | 0.13 (0.11:0.15), <0.001 | 0.13 (0.11:0.15), <0.001 | 0.11 (0.08:0.15), <0.001 | |
| **Year of birth** | | | | | -0.02 (-0.05:0.02), 0.290 | -0.11 (-0.13:-0.09), <0.001 |

**Supplementary Table 8. Linear regression models for Schizophrenia (SCZ) polygenic score in ADHD patients N=20,111**. Abbreviations: PC=principal component; Year of diagnosis (ADHD)=first recorded ADHD diagnosis for each individuals between 1994:2016 obtained from Danish national registers; Age at diagnosis (ADHD)=age at first recorded ADHD diagnosis for each individual between 1:35 obtained from Danish national health registers; Year of birth= between 1981:2008 obtained from Danish national health registers.

| **Predictor** | **Model 0** | **Model 1** | **Model 2** | **Model 3** | **Model 4** | **Model 5** |
|---|---|---|---|---|---|---|
| **(Intercept)** | 0.02 (-0.00:0.05), 0.081 | 0.07 (0.01:0.14), 0.019 | 0.02 (-0.04:0.09), 0.528 | -0.07 (-0.11:-0.02), 0.003 | 0.11 (0.00:0.21), 0.048 | 0.11 (0.07:0.15), <0.001 |
| **PC1** | 19.10 (14.40:23.80), <0.001 | 19.08 (14.38:23.78), <0.001 | 19.71 (15.01:24.41), <0.001 | 19.62 (14.92:24.33), <0.001 | 19.71 (15.01:24.41), <0.001 | 19.70 (15.00:24.40), <0.001 |
| **PC2** | 3.47 (-1.05:8.00), 0.133 | 3.51 (-1.02:8.03), 0.129 | 3.60 (-0.92:8.12), 0.119 | 3.51 (-1.01:8.04), 0.128 | 3.60 (-0.92:8.13), 0.118 | 3.61 (-0.91:8.13), 0.118 |
| **PC3** | -3.81 (-8.53:0.91), 0.114 | -3.89 (-8.61:0.83), 0.106 | -3.92 (-8.63:0.80), 0.103 | -3.76 (-8.48:0.96), 0.118 | -3.91 (-8.63:0.80), 0.104 | -3.92 (-8.63:0.80), 0.104 |
| **PC4** | 0.58 (-3.96:5.13), 0.802 | 0.57 (-3.97:5.12), 0.804 | 0.43 (-4.11:4.98), 0.851 | 0.48 (-4.06:5.02), 0.837 | 0.43 (-4.11:4.97), 0.853 | 0.43 (-4.11:4.97), 0.853 |
| **PC5** | -3.05 (-7.60:1.50), 0.189 | -3.01 (-7.56:1.54), 0.195 | -3.07 (-7.62:1.47), 0.185 | -3.14 (-7.68:1.41), 0.176 | -3.08 (-7.62:1.47), 0.185 | -3.07 (-7.62:1.47), 0.185 |
| **PC6** | 9.43 (4.91:13.96), <0.001 | 9.48 (4.96:14.01), <0.001 | 9.36 (4.84:13.89), <0.001 | 9.29 (4.77:13.82), <0.001 | 9.36 (4.83:13.88), <0.001 | 9.36 (4.84:13.88), <0.001 |
| **PC7** | 14.42 (9.89:18.96), <0.001 | 14.44 (9.91:18.97), <0.001 | 14.39 (9.86:18.92), <0.001 | 14.37 (9.84:18.90), <0.001 | 14.40 (9.87:18.93), <0.001 | 14.40 (9.87:18.93), <0.001 |
| **PC8** | 1.90 (-2.62:6.43), 0.410 | 1.91 (-2.62:6.44), 0.408 | 1.93 (-2.59:6.45), 0.403 | 1.91 (-2.61:6.44), 0.407 | 1.92 (-2.60:6.45), 0.405 | 1.92 (-2.60:6.45), 0.405 |
| **PC9** | -10.75 (-15.32:-6.18), <0.001 | -10.76 (-15.33:-6.19), <0.001 | -10.68 (-15.25:-6.12), <0.001 | -10.67 (-15.24:-6.11), <0.001 | -10.68 (-15.25:-6.12), <0.001 | -10.68 (-15.25:-6.12), <0.001 |
| **PC10** | -0.51 (-5.08:4.05), 0.825 | -0.52 (-5.09:4.04), 0.823 | -0.53 (-5.08:4.03), 0.821 | -0.51 (-5.07:4.05), 0.826 | -0.52 (-5.07:4.04), 0.825 | -0.52 (-5.07:4.04), 0.825 |
| **Sex** | -0.03 (-0.06:0.00), 0.084 | -0.03 (-0.06:-0.00), 0.048 | -0.02 (-0.05:0.02), 0.318 | -0.01 (-0.04:0.02), 0.491 | -0.02 (-0.05:0.02), 0.324 | -0.02 (-0.05:0.01), 0.305 |
| **Year of diagnosis (ADHD)** | | -0.03 (-0.06:0.00), 0.074 | -0.06 (-0.10:-0.03), <0.001 | | | |
| **Age at diagnosis (ADHD)** | | | 0.07 (0.04:0.09), <0.001 | 0.05 (0.03:0.07), <0.001 | 0.00 (-0.03:0.04), 0.912 | |
| **Year of birth** | | | | | -0.06 (-0.10:-0.03), <0.001 | -0.07 (-0.09:-0.04), <0.001 |

**Supplementary Table 9. Linear regression models for Addiction polygenic score in ADHD patients N=20,111**. Abbreviations: PC=principal component; Year of diagnosis (ADHD)=first recorded ADHD diagnosis for each individuals between 1994:2016 obtained from Danish national registers; Age at diagnosis (ADHD)=age at first recorded ADHD diagnosis for each individual between 1:35 obtained from Danish national health registers; Year of birth= between 1981:2008 obtained from Danish national health registers.

| Predictors | Model 0 | Model 1 | Model 2 | Model 3 | Model 4 | Model 5 |
|---|---|---|---|---|---|---|
| **(Intercept)** | 0.29 (0.26:0.31), p<0.001 | 0.23 (0.17:0.29), p<0.001 | 0.13 (0.06:0.19), p<0.001 | 0.07 (0.03:0.12), p=0.001 | 0.17 (0.07:0.28), p=0.001 | 0.44 (0.40:0.48), p<0.001 |
| **PC1** | 7.44 (2.74:12.15), p=0.002 | 7.46 (2.76:12.17), p=0.002 | 8.74 (4.04:13.43), p<0.001 | 8.68 (3.99:13.38), p<0.001 | 8.73 (4.04:13.43), p<0.001 | 8.49 (3.80:13.19), p<0.001 |
| **PC2** | -3.53 (-8.06:1.00), p=0.127 | -3.56 (-8.09:0.97), p=0.123 | -3.38 (-7.89:1.14), p=0.143 | -3.42 (-7.94:1.09), p=0.137 | -3.37 (-7.89:1.14), p=0.143 | -3.29 (-7.81:1.23), p=0.153 |
| **PC3** | -5.30 (-10.03:-0.58), p=0.028 | -5.22 (-9.95:-0.50), p=0.030 | -5.28 (-9.99:-0.58), p=0.028 | -5.19 (-9.90:-0.48), p=0.031 | -5.28 (-9.99:-0.57), p=0.028 | -5.49 (-10.20:-0.78), p=0.022 |
| **PC4** | 0.04 (-4.51:4.59), p=0.985 | 0.05 (-4.50:4.60), p=0.982 | -0.23 (-4.76:4.31), p=0.921 | -0.20 (-4.74:4.33), p=0.930 | -0.23 (-4.77:4.30), p=0.920 | -0.22 (-4.76:4.31), p=0.923 |
| **PC5** | -0.70 (-5.26:3.85), p=0.762 | -0.74 (-5.30:3.81), p=0.749 | -0.88 (-5.42:3.66), p=0.705 | -0.91 (-5.45:3.63), p=0.693 | -0.88 (-5.42:3.66), p=0.704 | -0.75 (-5.29:3.79), p=0.746 |
| **PC6** | 9.27 (4.74:13.80), p<0.001 | 9.22 (4.68:13.75), p<0.001 | 8.97 (4.46:13.49), p<0.001 | 8.93 (4.42:13.45), p<0.001 | 8.97 (4.45:13.49), p<0.001 | 9.14 (4.62:13.66), p<0.001 |
| **PC7** | 9.51 (4.97:14.05), p<0.001 | 9.49 (4.95:14.03), p<0.001 | 9.39 (4.87:13.92), p<0.001 | 9.38 (4.86:13.90), p<0.001 | 9.40 (4.87:13.92), p<0.001 | 9.47 (4.94:14.00), p<0.001 |
| **PC8** | 3.42 (-1.12:7.95), p=0.139 | 3.41 (-1.12:7.94), p=0.140 | 3.45 (-1.06:7.97), p=0.134 | 3.44 (-1.07:7.96), p=0.135 | 3.45 (-1.07:7.97), p=0.135 | 3.45 (-1.07:7.97), p=0.134 |
| **PC9** | -4.61 (-9.18:-0.04), p=0.048 | -4.60 (-9.17:-0.02), p=0.049 | -4.44 (-8.99:0.12), p=0.056 | -4.43 (-8.99:0.13), p=0.057 | -4.44 (-8.99:0.12), p=0.056 | -4.50 (-9.06:-0.07), p=0.053 |
| **PC10** | 0.56 (-4.01:5.13), p=0.810 | 0.57 (-4.00:5.14), p=0.808 | 0.56 (-3.99:5.12), p=0.809 | 0.57 (-3.98:5.12), p=0.806 | 0.57 (-3.99:5.12), p=0.807 | 0.56 (-4.00:5.12), p=0.810 |
| **Sex** | -0.04 (-0.07:-0.01), p=0.010 | -0.04 (-0.07:-0.00), p=0.023 | -0.01 (-0.04:0.03), p=0.731 | -0.00 (-0.03:0.03), p=0.870 | -0.01 (-0.04:0.03), p=0.737 | -0.02 (-0.05:0.01), p=0.172 |
| **Year of diagnosis (ADHD)** | | 0.03 (-0.00:0.06), p=0.067 | -0.04 (-0.07:-0.00), p=0.037 | | | |
| **Age at diagnosis (ADHD)** | | | 0.13 (0.11:0.15), p<0.001 | 0.12 (0.10:0.14), p<0.001 | 0.09 (0.06:0.13), p<0.001 | |
| **Year of birth** | | | | | -0.04 (-0.07:-0.00), p=0.040 | -0.11 (-0.13:-0.09), p<0.001 |

**Supplementary Table 10. Linear regression models for Education polygenic score in ADHD patients N=20,111.** Abbreviations: PC=principal component; Year of diagnosis (ADHD)=first recorded ADHD diagnosis for each individuals between 1994:2016 obtained from Danish national registers; Age at diagnosis (ADHD)=age at first recorded ADHD diagnosis for each individual between 1:35 obtained from Danish national health registers; Year of birth= between 1981:2008 obtained from Danish national health registers.

| Predictors | Model 0 | Model 1 | Model 2 | Model 3 | Model 4 | Model 5 |
|---|---|---|---|---|---|---|
| **(Intercept)** | -0.43 (-0.46:-0.41), p<0.001 | -0.31 (-0.37:-0.25), p<0.001 | -0.24 (-0.31:-0.18), p<0.001 | -0.28 (-0.33:-0.24), p<0.001 | -0.21 (-0.32:-0.11), p<0.001 | -0.52 (-0.56:-0.48), p<0.001 |
| **PC1** | -27.04 (-31.66:-22.42), p<0.001 | -27.09 (-31.71:-22.46), p<0.001 | -27.89 (-32.51:-23.27), p<0.001 | -27.93 (-32.55:-23.31), p<0.001 | -27.89 (-32.51:-23.27), p<0.001 | -27.62 (-32.24:-23.00), p<0.001 |
| **PC2** | 2.48 (-1.98:6.93), p=0.276 | 2.56 (-1.89:7.01), p=0.260 | 2.44 (-2.00:6.88), p=0.282 | 2.40 (-2.04:6.85), p=0.289 | 2.44 (-2.00:6.88), p=0.282 | 2.35 (-2.10:6.79), p=0.301 |
| **PC3** | 0.06 (-4.58:4.70), p=0.980 | -0.13 (-4.77:4.51), p=0.956 | -0.09 (-4.73:4.54), p=0.969 | -0.02 (-4.65:4.61), p=0.993 | -0.08 (-4.71:4.55), p=0.973 | 0.16 (-4.47:4.80), p=0.945 |
| **PC4** | -2.53 (-7.00:1.94), p=0.267 | -2.55 (-7.02:1.92), p=0.263 | -2.38 (-6.84:2.09), p=0.297 | -2.36 (-6.82:2.11), p=0.301 | -2.38 (-6.84:2.09), p=0.297 | -2.39 (-6.85:2.08), p=0.295 |
| **PC5** | -1.15 (-5.63:3.32), p=0.614 | -1.06 (-5.53:3.42), p=0.643 | -0.97 (-5.44:3.49), p=0.670 | -1.00 (-5.47:3.47), p=0.660 | -0.98 (-5.44:3.49), p=0.668 | -1.13 (-5.60:3.35), p=0.622 |
| **PC6** | 4.85 (0.40:9.30), p=0.033 | 4.97 (0.52:9.42), p=0.029 | 5.12 (0.68:9.57), p=0.024 | 5.09 (0.64:9.54), p=0.025 | 5.12 (0.67:9.56), p=0.024 | 4.92 (0.47:9.37), p=0.030 |
| **PC7** | 1.55 (-2.91:6.01), p=0.497 | 1.59 (-2.87:6.04), p=0.485 | 1.65 (-2.80:6.10), p=0.467 | 1.64 (-2.81:6.09), p=0.470 | 1.65 (-2.80:6.10), p=0.467 | 1.57 (-2.89:6.03), p=0.490 |
| **PC8** | 4.13 (-0.32:8.59), p=0.069 | 4.15 (-0.30:8.60), p=0.068 | 4.12 (-0.32:8.57), p=0.069 | 4.12 (-0.33:8.56), p=0.070 | 4.12 (-0.33:8.56), p=0.069 | 4.11 (-0.34:8.56), p=0.070 |
| **PC9** | -8.81 (-13.30:-4.32), p<0.001 | -8.84 (-13.33:-4.35), p<0.001 | -8.94 (-13.43:-4.46), p<0.001 | -8.94 (-13.42:-4.45), p<0.001 | -8.94 (-13.43:-4.46), p<0.001 | -8.87 (-13.36:-4.38), p<0.001 |
| **PC10** | 2.54 (-1.95:7.03), p=0.267 | 2.52 (-1.96:7.01), p=0.270 | 2.53 (-1.95:7.01), p=0.269 | 2.53 (-1.95:7.02), p=0.268 | 2.53 (-1.95:7.01), p=0.268 | 2.54 (-1.94:7.03), p=0.267 |
| **Sex** | 0.03 (-0.00:0.06), p=0.079 | 0.02 (-0.01:0.05), p=0.276 | -0.00 (-0.03:0.03), p=0.887 | 0.00 (-0.03:0.03), p=0.997 | -0.00 (-0.03:0.03), p=0.905 | 0.02 (-0.01:0.05), p=0.284 |
| **Year of diagnosis (ADHD)** | | -0.07 (-0.11:-0.04), p<0.001 | -0.03 (-0.06:0.00), p=0.093 | | | - |
| **Age at diagnosis (ADHD)** | | | -0.08 (-0.10:-0.06), p<0.001 | -0.09 (-0.11:-0.07), p<0.001 | -0.11 (-0.14:-0.07), p<0.001 | - |
| **Year of birth** | | | | | -0.03 (-0.06:0.01), p=0.148 | 0.06 (0.04:0.08), p<0.001 |

**Supplementary Table 11. Linear regression models for Intelligence polygenic score in ADHD patients N=20,111**. Abbreviations: PC=principal component; Year of diagnosis (ADHD)=first recorded ADHD diagnosis for each individuals between 1994:2016 obtained from Danish national registers; Age at diagnosis (ADHD)=age at first recorded ADHD diagnosis for each individual between 1:35 obtained from Danish national health registers; Year of birth= between 1981:2008 obtained from Danish national health registers.

| Predictor | Model 0 | Model 1 | Model 2 | Model 3 | Model 4 | Model 5 |
|---|---|---|---|---|---|---|
| **(Intercept)** | -0.27 (-0.30:-0.25), <0.001 | -0.23 (-0.29:-0.17), <0.001 | -0.20 (-0.26:-0.14), <0.001 | -0.21 (-0.25:-0.16), <0.001 | -0.20 (-0.30:-0.10), <0.001 | -0.31 (-0.35:-0.27), <0.001 |
| **PC1** | -20.13 (-24.75:-15.50), <0.001 | -20.14 (-24.77:-15.52), <0.001 | -20.50 (-25.13:-15.87), <0.001 | -20.51 (-25.14:-15.88), <0.001 | -20.51 (-25.14:-15.88), <0.001 | -20.41 (-25.04:-15.78), <0.001 |
| **PC2** | 0.46 (-3.99:4.91), 0.840 | 0.49 (-3.97:4.94), 0.830 | 0.43 (-4.02:4.89), 0.849 | 0.43 (-4.02:4.88), 0.851 | 0.43 (-4.02:4.88), 0.850 | 0.40 (-4.06:4.85), 0.861 |
| **PC3** | -1.07 (-5.72:3.57), 0.651 | -1.14 (-5.78:3.51), 0.632 | -1.12 (-5.76:3.53), 0.637 | -1.11 (-5.75:3.54), 0.640 | -1.11 (-5.75:3.53), 0.639 | -1.02 (-5.67:3.62), 0.666 |
| **PC4** | -4.59 (-9.07:-0.12), 0.044 | -4.60 (-9.07:-0.13), 0.044 | -4.52 (-8.99:-0.05), 0.048 | -4.52 (-8.99:-0.04), 0.048 | -4.52 (-8.99:-0.04), 0.048 | -4.52 (-8.99:-0.05), 0.048 |
| **PC5** | -6.19 (-10.67:-1.71), 0.007 | -6.16 (-10.64:-1.68), 0.007 | -6.12 (-10.60:-1.65), 0.007 | -6.13 (-10.60:-1.65), 0.007 | -6.13 (-10.60:-1.65), 0.007 | -6.18 (-10.66:-1.70), 0.007 |
| **PC6** | 4.39 (-0.07:8.84), 0.054 | 4.42 (-0.03:8.88), 0.052 | 4.49 (0.04:8.95), 0.048 | 4.49 (0.03:8.94), 0.048 | 4.49 (0.04:8.95), 0.048 | 4.42 (-0.04:8.88), 0.052 |
| **PC7** | -4.00 (-8.46:0.46), 0.079 | -3.99 (-8.45:0.48), 0.080 | -3.96 (-8.42:0.50), 0.082 | -3.96 (-8.42:0.50), 0.082 | -3.96 (-8.42:0.50), 0.082 | -3.99 (-8.45:0.47), 0.080 |
| **PC8** | 9.03 (4.57:13.48), <0.001 | 9.03 (4.58:13.49), <0.001 | 9.02 (4.56:13.47), <0.001 | 9.02 (4.56:13.47), <0.001 | 9.02 (4.56:13.47), <0.001 | 9.02 (4.56:13.47), <0.001 |
| **PC9** | -1.85 (-6.35:2.64), 0.420 | -1.86 (-6.36:2.63), 0.417 | -1.91 (-6.40:2.59), 0.406 | -1.91 (-6.40:2.59), 0.406 | -1.91 (-6.40:2.59), 0.406 | -1.88 (-6.38:2.61), 0.412 |
| **PC10** | 0.84 (-3.65:5.33), 0.715 | 0.83 (-3.66:5.32), 0.717 | 0.83 (-3.66:5.32), 0.716 | 0.83 (-3.66:5.32), 0.716 | 0.83 (-3.66:5.32), 0.716 | 0.84 (-3.65:5.33), 0.715 |
| **Sex** | 0.03 (-0.00:0.06), 0.091 | 0.02 (-0.01:0.05), 0.145 | 0.01 (-0.02:0.04), 0.374 | 0.01 (-0.02:0.04), 0.360 | 0.01 (-0.02:0.04), 0.366 | 0.02 (-0.01:0.05), 0.176 |
| **Year of diagnosis (ADHD)** | | -0.02 (-0.06:0.01), 0.152 | -0.00 (-0.04:0.03), 0.801 | | | |
| **Age at diagnosis (ADHD)** | | | -0.04 (-0.06:-0.02), 0.001 | -0.04 (-0.06:-0.02), <0.001 | -0.04 (-0.07:-0.01), 0.023 | |
| **Year of birth** | | | | | -0.00 (-0.04:0.03), 0.929 | 0.03 (0.01:0.05), 0.004 |

**Supplementary Table 12. Linear regression models for Risk-taking polygenic score in ADHD patients N=20,111**. Abbreviations: PC=principal component; Year of diagnosis (ADHD)=first recorded ADHD diagnosis for each individuals between 1994:2016 obtained from Danish national registers; Age at diagnosis (ADHD)=age at first recorded ADHD diagnosis for each individual between 1:35 obtained from Danish national health registers; Year of birth= between 1981:2008 obtained from Danish national health registers.

| Predictor | Model 0 | Model 1 | Model 2 | Model 3 | Model 4 | Model 5 |
|---|---|---|---|---|---|---|
| **(Intercept)** | 0.22 (0.20:0.25), <0.001 | 0.24 (0.18:0.30), <0.001 | 0.22 (0.15:0.28), <0.001 | 0.19 (0.14:0.23), <0.001 | 0.26 (0.16:0.36), <0.001 | 0.26 (0.22:0.30), <0.001 |
| **PC1** | 3.39 (-1.30:8.08), 0.157 | 3.38 (-1.31:8.07), 0.158 | 3.64 (-1.06:8.34), 0.129 | 3.61 (-1.09:8.30), 0.132 | 3.64 (-1.05:8.34), 0.128 | 3.64 (-1.05:8.34), 0.128 |
| **PC2** | 4.98 (0.46:9.50), 0.031 | 4.99 (0.48:9.51), 0.030 | 5.03 (0.51:9.55), 0.029 | 5.00 (0.48:9.51), 0.030 | 5.04 (0.52:9.55), 0.029 | 5.04 (0.52:9.55), 0.029 |
| **PC3** | -0.70 (-5.41:4.01), 0.770 | -0.73 (-5.44:3.98), 0.761 | -0.74 (-5.45:3.97), 0.757 | -0.68 (-5.39:4.03), 0.776 | -0.75 (-5.46:3.96), 0.755 | -0.75 (-5.46:3.96), 0.755 |
| **PC4** | 0.42 (-4.12:4.95), 0.857 | 0.41 (-4.12:4.95), 0.858 | 0.36 (-4.18:4.89), 0.877 | 0.37 (-4.16:4.91), 0.872 | 0.35 (-4.18:4.89), 0.879 | 0.35 (-4.18:4.89), 0.879 |
| **PC5** | 1.03 (-3.51:5.57), 0.656 | 1.05 (-3.49:5.59), 0.652 | 1.02 (-3.52:5.56), 0.660 | 1.00 (-3.55:5.54), 0.667 | 1.02 (-3.52:5.56), 0.659 | 1.02 (-3.52:5.56), 0.659 |
| **PC6** | 7.79 (3.27:12.31), 0.001 | 7.81 (3.29:12.33), 0.001 | 7.76 (3.24:12.28), 0.001 | 7.73 (3.21:12.25), 0.001 | 7.76 (3.24:12.28), 0.001 | 7.76 (3.24:12.28), 0.001 |
| **PC7** | -2.58 (-7.11:1.94), 0.264 | -2.58 (-7.10:1.95), 0.265 | -2.60 (-7.12:1.93), 0.261 | -2.60 (-7.13:1.92), 0.259 | -2.59 (-7.12:1.93), 0.262 | -2.59 (-7.12:1.93), 0.262 |
| **PC8** | -1.26 (-5.78:3.26), 0.586 | -1.25 (-5.77:3.26), 0.586 | -1.25 (-5.76:3.27), 0.589 | -1.25 (-5.77:3.27), 0.587 | -1.25 (-5.77:3.27), 0.588 | -1.25 (-5.77:3.27), 0.588 |
| **PC9** | -3.98 (-8.54:0.58), 0.087 | -3.98 (-8.54:0.58), 0.087 | -3.95 (-8.51:0.61), 0.089 | -3.95 (-8.51:0.61), 0.090 | -3.95 (-8.51:0.61), 0.089 | -3.95 (-8.51:0.61), 0.089 |
| **PC10** | -4.12 (-8.68:0.43), 0.076 | -4.12 (-8.68:0.43), 0.076 | -4.13 (-8.68:0.43), 0.076 | -4.12 (-8.68:0.43), 0.076 | -4.12 (-8.68:0.43), 0.076 | -4.12 (-8.68:0.43), 0.076 |
| **Sex** | -0.06 (-0.09:-0.03), <0.001 | -0.07 (-0.10:-0.03), <0.001 | -0.06 (-0.09:-0.03), <0.001 | -0.06 (-0.09:-0.03), <0.001 | -0.06 (-0.09:-0.03), <0.001 | -0.06 (-0.09:-0.03), <0.001 |
| **Year of diagnosis (ADHD)** | | -0.01 (-0.04:0.02), 0.524 | -0.02 (-0.06:0.01), 0.167 | | | |
| **Age at diagnosis (ADHD)** | | | 0.03 (0.00:0.05), 0.017 | 0.02 (0.00:0.04), 0.039 | -0.00 (-0.04:0.03), 0.984 | |
| **Year of birth** | | | | | -0.03 (-0.06:0.01), 0.119 | -0.03 (-0.05:-0.01), 0.010 |

**Supplementary Table 13. Linear regression models for Neuroticism polygenic score in ADHD patients N=20,111**. Abbreviations: PC=principal component; Year of diagnosis (ADHD)=first recorded ADHD diagnosis for each individuals between 1994:2016 obtained from Danish national registers; Age at diagnosis (ADHD)=age at first recorded ADHD diagnosis for each individual between 1:35 obtained from Danish national health registers; Year of birth= between 1981:2008 obtained from Danish national health registers.

| Predictor | Model 0 | Model 1 | Model 2 | Model 3 | Model 4 | Model 5 |
|---|---|---|---|---|---|---|
| **(Intercept)** | 0.19 (0.16:0.21), <0.001 | 0.12 (0.06:0.18), <0.001 | 0.05 (-0.01:0.12), 0.115 | 0.04 (0.00:0.09), 0.046 | 0.06 (-0.05:0.16), 0.291 | 0.29 (0.25:0.32), <0.001 |
| **PC1** | 4.91 (0.26:9.56), 0.038 | 4.93 (0.29:9.58), 0.037 | 5.77 (1.13:10.42), 0.015 | 5.77 (1.12:10.41), 0.015 | 5.77 (1.13:10.42), 0.015 | 5.56 (0.92:10.21), 0.019 |
| **PC2** | -4.43 (-8.91:-0.04), 0.052 | -4.48 (-8.95:-0.00), 0.050 | -4.36 (-8.82:-0.11), 0.056 | -4.36 (-8.83:-0.10), 0.056 | -4.36 (-8.82:-0.11), 0.056 | -4.29 (-8.76:-0.18), 0.060 |
| **PC3** | -4.89 (-9.55:-0.22), 0.040 | -4.79 (-9.45:-0.12), 0.044 | -4.83 (-9.49:-0.17), 0.042 | -4.81 (-9.47:-0.15), 0.043 | -4.82 (-9.48:-0.16), 0.043 | -5.01 (-9.67:-0.34), 0.035 |
| **PC4** | 2.50 (-1.99:7.00), 0.275 | 2.51 (-1.98:7.01), 0.273 | 2.33 (-2.16:6.81), 0.310 | 2.33 (-2.16:6.82), 0.309 | 2.33 (-2.16:6.81), 0.310 | 2.33 (-2.16:6.82), 0.308 |
| **PC5** | 6.41 (1.91:10.90), 0.005 | 6.35 (1.86:10.83), 0.006 | 6.27 (1.78:10.76), 0.006 | 6.26 (1.77:10.75), 0.006 | 6.27 (1.77:10.76), 0.006 | 6.38 (1.88:10.87), 0.005 |
| **PC6** | -11.05 (-15.52:-6.57), <0.001 | -11.11 (-15.59:-6.63), <0.001 | -11.27 (-15.74:-6.80), <0.001 | -11.28 (-15.75:-6.81), <0.001 | -11.27 (-15.74:-6.80), <0.001 | -11.13 (-15.60:-6.65), <0.001 |
| **PC7** | -5.11 (-9.59:-0.62), 0.026 | -5.13 (-9.61:-0.65), 0.025 | -5.20 (-9.67:-0.72), 0.023 | -5.20 (-9.67:-0.72), 0.023 | -5.20 (-9.67:-0.72), 0.023 | -5.13 (-9.61:-0.66), 0.025 |
| **PC8** | -21.38 (-25.86:-16.91), <0.001 | -21.39 (-25.87:-16.92), <0.001 | -21.37 (-25.83:-16.90), <0.001 | -21.37 (-25.84:-16.90), <0.001 | -21.37 (-25.84:-16.90), <0.001 | -21.36 (-25.83:-16.89), <0.001 |
| **PC9** | 9.30 (4.78:13.81), <0.001 | 9.31 (4.80:13.83), <0.001 | 9.42 (4.91:13.93), <0.001 | 9.42 (4.91:13.93), <0.001 | 9.42 (4.91:13.93), <0.001 | 9.37 (4.86:13.88), <0.001 |
| **PC10** | 4.19 (-0.32:8.71), 0.069 | 4.20 (-0.31:8.71), 0.068 | 4.20 (-0.31:8.70), 0.068 | 4.20 (-0.31:8.70), 0.068 | 4.20 (-0.31:8.70), 0.068 | 4.19 (-0.32:8.70), 0.068 |
| **Sex** | -0.05 (-0.08:-0.02), 0.002 | -0.04 (-0.07:-0.01), 0.006 | -0.02 (-0.05:0.01), 0.156 | -0.02 (-0.05:0.01), 0.162 | -0.02 (-0.05:0.01), 0.158 | -0.04 (-0.07:-0.01), 0.020 |
| **Year of diagnosis (ADHD)** | | 0.04 (0.01:0.07), 0.018 | -0.01 (-0.04:0.03), 0.752 | | | |
| **Age at diagnosis (ADHD)** | | | 0.09 (0.07:0.11), <0.001 | 0.09 (0.07:0.11), <0.001 | 0.08 (0.05:0.12), <0.001 | |
| **Year of birth** | | | | | -0.00 (-0.04:0.03), 0.807 | -0.07 (-0.09:-0.05), <0.001 |

**Supplementary Table 14. Linear regression models for ASD polygenic score in ASD patients N=17,071**. Abbreviations: PC=principal component; Year of diagnosis (ADHD)=first recorded ADHD diagnosis for each individuals between 1994:2016 obtained from Danish national registers; Age at diagnosis (ADHD)=age at first recorded ADHD diagnosis for each individual between 1:35 obtained from Danish national health registers; Year of birth= between 1981:2008 obtained from Danish national health registers.

| Predictor | Model 0 | Model 1 | Model 2 | Model 3 | Model 4 | Model 5 |
|---|---|---|---|---|---|---|
| **(Intercept)** | 0.31 (0.28:0.34), <0.001 | 0.43 (0.37:0.48), <0.001 | 0.44 (0.38:0.50), <0.001 | 0.35 (0.30:0.40), <0.001 | 0.53 (0.44:0.62), <0.001 | 0.35 (0.30:0.40), <0.001 |
| **PC1** | 12.15 (7.12:17.19), <0.001 | 12.11 (7.07:17.14), <0.001 | 11.97 (6.93:17.02), <0.001 | 11.73 (6.68:16.77), <0.001 | 11.97 (6.92:17.01), <0.001 | 12.46 (7.42:17.51), <0.001 |
| **PC2** | 8.21 (3.29:13.14), 0.001 | 8.25 (3.33:13.17), 0.001 | 8.24 (3.32:13.16), 0.001 | 8.21 (3.29:13.13), 0.001 | 8.23 (3.31:13.15), 0.001 | 8.22 (3.30:13.15), 0.001 |
| **PC3** | -7.17 (-12.27:-2.08), 0.006 | -7.45 (-12.54:-2.37), 0.004 | -7.54 (-12.64:-2.45), 0.004 | -7.50 (-12.60:-2.40), 0.004 | -7.52 (-12.61:-2.43), 0.004 | -7.02 (-12.11:-1.92), 0.007 |
| **PC4** | 2.87 (-2.07:7.81), 0.255 | 2.74 (-2.20:7.67), 0.278 | 2.75 (-2.19:7.69), 0.275 | 2.90 (-2.04:7.85), 0.250 | 2.76 (-2.18:7.70), 0.274 | 2.80 (-2.15:7.74), 0.268 |
| **PC5** | -3.31 (-8.28:1.65), 0.191 | -3.25 (-8.21:1.71), 0.199 | -3.23 (-8.19:1.74), 0.202 | -3.24 (-8.20:1.73), 0.202 | -3.24 (-8.21:1.72), 0.200 | -3.36 (-8.32:1.61), 0.185 |
| **PC6** | 5.40 (0.43:10.37), 0.033 | 5.43 (0.47:10.40), 0.032 | 5.47 (0.50:10.43), 0.031 | 5.52 (0.55:10.48), 0.030 | 5.49 (0.53:10.46), 0.030 | 5.33 (0.37:10.30), 0.035 |
| **PC7** | -9.59 (-14.51:-4.67), <0.001 | -9.63 (-14.54:-4.71), <0.001 | -9.60 (-14.52:-4.68), <0.001 | -9.50 (-14.42:-4.58), <0.001 | -9.59 (-14.50:-4.67), <0.001 | -9.67 (-14.59:-4.75), <0.001 |
| **PC8** | -4.28 (-9.23:0.67), 0.090 | -4.33 (-9.27:0.62), 0.087 | -4.33 (-9.28:0.62), 0.086 | -4.30 (-9.25:0.65), 0.089 | -4.32 (-9.27:0.63), 0.087 | -4.29 (-9.24:0.67), 0.090 |
| **PC9** | 7.69 (2.74:12.64), 0.002 | 7.65 (2.71:12.60), 0.002 | 7.67 (2.72:12.62), 0.002 | 7.74 (2.79:12.69), 0.002 | 7.66 (2.72:12.61), 0.002 | 7.64 (2.69:12.59), 0.002 |
| **PC10** | 5.48 (0.55:10.40), 0.029 | 5.30 (0.38:10.22), 0.035 | 5.31 (0.39:10.23), 0.034 | 5.47 (0.55:10.39), 0.029 | 5.32 (0.40:10.24), 0.034 | 5.42 (0.50:10.34), 0.031 |
| **Sex** | 0.04 (0.01:0.08), 0.012 | 0.03 (-0.00:0.07), 0.068 | 0.03 (-0.00:0.07), 0.085 | 0.04 (0.00:0.07), 0.036 | 0.03 (-0.00:0.07), 0.084 | 0.05 (0.01:0.08), 0.010 |
| **Year of diagnosis (ASD)** | | -0.08 (-0.10:-0.05), <0.001 | -0.07 (-0.10:-0.04), <0.001 | | | |
| **Age at diagnosis (ASD)** | | | -0.01 (-0.04:0.02), 0.460 | -0.03 (-0.06:-0.01), 0.013 | -0.08 (-0.12:-0.05), <0.001 | |
| **Year of birth** | | | | | -0.07 (-0.10:-0.04), <0.001 | -0.03 (-0.05:-0.00), 0.034 |

**Supplementary Table 15. Linear regression models for ADHD polygenic score in ASD patients N=17,071**. Abbreviations: PC=principal component; Year of diagnosis (ADHD)=first recorded ADHD diagnosis for each individuals between 1994:2016 obtained from Danish national registers; Age at diagnosis (ADHD)=age at first recorded ADHD diagnosis for each individual between 1:35 obtained from Danish national health registers; Year of birth= between 1981:2008 obtained from Danish national health registers.

| Predictor | Model 0 | Model 1 | Model 2 | Model 3 | Model 4 | Model 5 |
|---|---|---|---|---|---|---|
| **(Intercept)** | 0.03 (-0.00:0.06), 0.084 | -0.04 (-0.10:0.01), 0.115 | -0.04 (-0.10:0.01), 0.141 | 0.01 (-0.04:0.05), 0.804 | -0.11 (-0.20:-0.02), 0.022 | -0.01 (-0.05:0.04), 0.831 |
| **PC1** | 13.07 (8.04:18.10), <0.001 | 13.10 (8.07:18.13), <0.001 | 13.11 (8.07:18.15), <0.001 | 13.26 (8.22:18.30), <0.001 | 13.11 (8.07:18.15), <0.001 | 12.83 (7.80:17.87), <0.001 |
| **PC2** | -2.67 (-7.59:2.24), 0.287 | -2.69 (-7.61:2.22), 0.283 | -2.69 (-7.61:2.22), 0.283 | -2.67 (-7.59:2.25), 0.287 | -2.68 (-7.60:2.23), 0.284 | -2.68 (-7.60:2.24), 0.285 |
| **PC3** | -8.46 (-13.55:-3.38), 0.001 | -8.30 (-13.38:-3.22), 0.001 | -8.30 (-13.39:-3.21), 0.001 | -8.32 (-13.41:-3.23), 0.001 | -8.31 (-13.40:-3.22), 0.001 | -8.59 (-13.67:-3.50), 0.001 |
| **PC4** | 3.79 (-1.14:8.73), 0.132 | 3.87 (-1.07:8.80), 0.124 | 3.87 (-1.07:8.80), 0.124 | 3.78 (-1.16:8.71), 0.134 | 3.87 (-1.07:8.80), 0.124 | 3.85 (-1.09:8.78), 0.127 |
| **PC5** | 0.98 (-3.98:5.94), 0.700 | 0.94 (-4.02:5.90), 0.711 | 0.94 (-4.02:5.90), 0.711 | 0.94 (-4.02:5.90), 0.710 | 0.95 (-4.01:5.91), 0.708 | 1.01 (-3.95:5.97), 0.690 |
| **PC6** | -0.39 (-5.35:4.57), 0.877 | -0.41 (-5.37:4.55), 0.871 | -0.41 (-5.37:4.55), 0.871 | -0.44 (-5.40:4.52), 0.861 | -0.43 (-5.39:4.53), 0.866 | -0.34 (-5.30:4.62), 0.893 |
| **PC7** | 8.59 (3.68:13.51), 0.001 | 8.62 (3.70:13.53), 0.001 | 8.61 (3.70:13.53), 0.001 | 8.56 (3.64:13.47), 0.001 | 8.61 (3.70:13.52), 0.001 | 8.65 (3.74:13.57), 0.001 |
| **PC8** | -5.16 (-10.10:-0.21), 0.041 | -5.13 (-10.07:-0.19), 0.042 | -5.13 (-10.07:-0.19), 0.042 | -5.15 (-10.10:-0.21), 0.041 | -5.14 (-10.08:-0.19), 0.042 | -5.15 (-10.10:-0.21), 0.041 |
| **PC9** | 17.93 (12.99:22.87), <0.001 | 17.95 (13.01:22.90), <0.001 | 17.95 (13.01:22.90), <0.001 | 17.91 (12.97:22.85), <0.001 | 17.96 (13.02:22.90), <0.001 | 17.97 (13.03:22.92), <0.001 |
| **PC10** | 6.53 (1.62:11.44), 0.009 | 6.63 (1.72:11.55), 0.008 | 6.63 (1.72:11.55), 0.008 | 6.53 (1.62:11.45), 0.009 | 6.63 (1.72:11.54), 0.008 | 6.57 (1.66:11.49), 0.009 |
| **Sex** | 0.05 (0.01:0.08), 0.010 | 0.05 (0.02:0.09), 0.003 | 0.05 (0.02:0.09), 0.003 | 0.05 (0.01:0.08), 0.007 | 0.05 (0.02:0.09), 0.003 | 0.04 (0.01:0.08), 0.012 |
| **Year of diagnosis (ASD)** | | 0.04 (0.02:0.07), 0.003 | 0.04 (0.01:0.07), 0.005 | | | |
| **Age at diagnosis (ASD)** | | | 0.00 (-0.03:0.03), 0.968 | 0.02 (-0.01:0.04), 0.279 | 0.04 (0.01:0.08), 0.010 | |
| **Year of birth** | | | | | 0.04 (0.01:0.08), 0.004 | 0.02 (-0.00:0.04), 0.103 |

**Supplementary Table 16. Linear regression models for Bipolar (BP) polygenic score in ASD patients N=17,071**. Abbreviations: PC=principal component; Year of diagnosis (ADHD)=first recorded ADHD diagnosis for each individuals between 1994:2016 obtained from Danish national registers; Age at diagnosis (ADHD)=age at first recorded ADHD diagnosis for each individual between 1:35 obtained from Danish national health registers; Year of birth= between 1981:2008 obtained from Danish national health registers.

| Predictor | Model 0 | Model 1 | Model 2 | Model 3 | Model 4 | Model 5 |
|---|---|---|---|---|---|---|
| **(Intercept)** | 0.13 (0.10:0.16), p<0.001 | 0.20 (0.14:0.25), p<0.001 | 0.18 (0.11:0.24), p<0.001 | 0.11 (0.07:0.16), p<0.001 | 0.25 (0.16:0.34), p<0.001 | 0.19 (0.14:0.24), p<0.001 |
| **PC1** | 13.89 (8.81:18.97), p<0.001 | 13.86 (8.78:18.95), p<0.001 | 14.17 (9.08:19.27), p<0.001 | 13.99 (8.90:19.09), p<0.001 | 14.18 (9.08:19.27), p<0.001 | 14.34 (9.25:19.43), p<0.001 |
| **PC2** | -6.40 (-11.37:-1.43), p=0.012 | -6.38 (-11.35:-1.42), p=0.012 | -6.38 (-11.34:-1.41), p=0.012 | -6.40 (-11.37:-1.43), p=0.012 | -6.38 (-11.35:-1.42), p=0.012 | -6.39 (-11.35:-1.42), p=0.012 |
| **PC3** | -5.28 (-10.41:-0.14), p=0.044 | -5.44 (-10.57:-0.30), p=0.038 | -5.23 (-10.37:-0.09), p=0.046 | -5.20 (-10.34:-0.06), p=0.048 | -5.21 (-10.36:-0.07), p=0.047 | -5.04 (-10.18:0.09), p=0.054 |
| **PC4** | 4.43 (-0.56:9.41), p=0.082 | 4.35 (-0.64:9.34), p=0.087 | 4.31 (-0.68:9.30), p=0.090 | 4.42 (-0.57:9.40), p=0.083 | 4.31 (-0.68:9.29), p=0.091 | 4.32 (-0.67:9.31), p=0.090 |
| **PC5** | 3.92 (-1.09:8.93), p=0.125 | 3.96 (-1.05:8.97), p=0.122 | 3.91 (-1.10:8.92), p=0.126 | 3.90 (-1.11:8.92), p=0.127 | 3.90 (-1.11:8.91), p=0.127 | 3.86 (-1.15:8.87), p=0.131 |
| **PC6** | 5.67 (0.66:10.69), p=0.027 | 5.69 (0.68:10.70), p=0.026 | 5.61 (0.60:10.62), p=0.028 | 5.64 (0.63:10.66), p=0.027 | 5.63 (0.61:10.64), p=0.028 | 5.57 (0.56:10.59), p=0.029 |
| **PC7** | -0.45 (-5.41:4.52), p=0.860 | -0.47 (-5.43:4.50), p=0.853 | -0.54 (-5.50:4.43), p=0.832 | -0.47 (-5.43:4.50), p=0.854 | -0.53 (-5.49:4.43), p=0.834 | -0.56 (-5.52:4.41), p=0.826 |
| **PC8** | -3.75 (-8.75:1.24), p=0.141 | -3.78 (-8.77:1.22), p=0.138 | -3.77 (-8.77:1.22), p=0.139 | -3.75 (-8.75:1.25), p=0.141 | -3.77 (-8.76:1.22), p=0.139 | -3.76 (-8.75:1.24), p=0.140 |
| **PC9** | -5.59 (-10.59:-0.60), p=0.028 | -5.62 (-10.61:-0.62), p=0.028 | -5.66 (-10.65:-0.66), p=0.026 | -5.61 (-10.60:-0.61), p=0.028 | -5.66 (-10.66:-0.67), p=0.026 | -5.67 (-10.67:-0.68), p=0.026 |
| **PC10** | 3.78 (-1.18:8.75), p=0.135 | 3.69 (-1.28:8.65), p=0.146 | 3.67 (-1.30:8.63), p=0.148 | 3.79 (-1.18:8.75), p=0.135 | 3.67 (-1.29:8.63), p=0.147 | 3.70 (-1.26:8.67), p=0.144 |
| **Sex** | 0.02 (-0.01:0.06), p=0.245 | 0.01 (-0.02:0.05), p=0.440 | 0.02 (-0.02:0.05), p=0.333 | 0.02 (-0.01:0.06), p=0.215 | 0.02 (-0.02:0.05), p=0.336 | 0.02 (-0.01:0.06), p=0.210 |
| **Year of diagnosis (ASD)** | | -0.04 (-0.07:-0.01), p=0.004 | -0.05 (-0.08:-0.02), p=0.001 | | | |
| **Age at diagnosis (ASD)** | | | 0.03 (-0.00:0.05), p=0.089 | 0.01 (-0.02:0.04), p=0.552 | -0.03 (-0.06:0.01), p=0.117 | |
| **Year of birth** | | | | | -0.05 (-0.08:-0.02), p=0.001 | -0.04 (-0.06:-0.01), p=0.002 |

**Supplementary Table 17. Linear regression models for Major depressiv disorder (MDD) polygenic score in ASD patients N=17,071**. Abbreviations: PC=principal component; Year of diagnosis (ADHD)=first recorded ADHD diagnosis for each individuals between 1994:2016 obtained from Danish national registers; Age at diagnosis (ADHD)=age at first recorded ADHD diagnosis for each individual between 1:35 obtained from Danish national health registers; Year of birth= between 1981:2008 obtained from Danish national health registers.

| Predictor | Model 0 | Model 1 | Model 2 | Model 3 | Model 4 | Model 5 |
|---|---|---|---|---|---|---|
| **(Intercept)** | 0.26 (0.23:0.29), p<0.001 | 0.19 (0.14:0.25), p<0.001 | 0.12 (0.06:0.18), p<0.001 | 0.13 (0.08:0.17), p<0.001 | 0.11 (0.03:0.20), p=0.012 | 0.33 (0.29:0.38), p<0.001 |
| **PC1** | 14.23 (9.25:19.21), p<0.001 | 14.25 (9.27:19.23), p<0.001 | 15.38 (10.40:20.37), p<0.001 | 15.41 (10.42:20.39), p<0.001 | 15.39 (10.40:20.37), p<0.001 | 14.79 (9.81:19.78), p<0.001 |
| **PC2** | -1.09 (-5.96:3.78), p=0.661 | -1.11 (-5.97:3.76), p=0.656 | -1.08 (-5.95:3.78), p=0.663 | -1.08 (-5.94:3.78), p=0.664 | -1.08 (-5.94:3.78), p=0.663 | -1.07 (-5.94:3.80), p=0.666 |
| **PC3** | -2.82 (-7.85:2.22), p=0.273 | -2.67 (-7.70:2.37), p=0.299 | -1.92 (-6.95:3.12), p=0.456 | -1.92 (-6.95:3.11), p=0.455 | -1.92 (-6.95:3.12), p=0.455 | -2.53 (-7.56:2.51), p=0.325 |
| **PC4** | 0.78 (-4.11:5.67), p=0.755 | 0.85 (-4.04:5.74), p=0.733 | 0.70 (-4.18:5.58), p=0.779 | 0.68 (-4.20:5.57), p=0.783 | 0.69 (-4.19:5.58), p=0.780 | 0.65 (-4.24:5.54), p=0.795 |
| **PC5** | 4.31 (-0.61:9.22), p=0.086 | 4.27 (-0.64:9.18), p=0.088 | 4.09 (-0.81:9.00), p=0.102 | 4.09 (-0.81:9.00), p=0.102 | 4.09 (-0.81:9.00), p=0.102 | 4.23 (-0.68:9.14), p=0.092 |
| **PC6** | -2.88 (-7.80:2.03), p=0.250 | -2.90 (-7.81:2.02), p=0.248 | -3.19 (-8.10:1.72), p=0.202 | -3.20 (-8.11:1.71), p=0.202 | -3.20 (-8.10:1.71), p=0.202 | -3.00 (-7.92:1.91), p=0.231 |
| **PC7** | -1.50 (-6.37:3.36), p=0.545 | -1.48 (-6.35:3.38), p=0.551 | -1.73 (-6.59:3.13), p=0.484 | -1.74 (-6.60:3.12), p=0.482 | -1.74 (-6.60:3.12), p=0.483 | -1.64 (-6.51:3.22), p=0.508 |
| **PC8** | -6.40 (-11.30:-1.50), p=0.010 | -6.38 (-11.27:-1.48), p=0.011 | -6.36 (-11.25:-1.47), p=0.011 | -6.36 (-11.25:-1.47), p=0.011 | -6.36 (-11.25:-1.47), p=0.011 | -6.40 (-11.30:-1.51), p=0.010 |
| **PC9** | 0.48 (-4.42:5.37), p=0.848 | 0.50 (-4.40:5.40), p=0.841 | 0.36 (-4.53:5.25), p=0.886 | 0.35 (-4.54:5.24), p=0.888 | 0.36 (-4.53:5.25), p=0.887 | 0.38 (-4.51:5.28), p=0.878 |
| **PC10** | 3.96 (-0.91:8.82), p=0.111 | 4.05 (-0.82:8.92), p=0.103 | 3.98 (-0.88:8.84), p=0.108 | 3.97 (-0.89:8.83), p=0.110 | 3.98 (-0.88:8.84), p=0.109 | 3.86 (-1.01:8.72), p=0.120 |
| **Sex** | -0.05 (-0.08:-0.01), p=0.007 | -0.04 (-0.08:-0.01), p=0.020 | -0.03 (-0.06:0.01), p=0.119 | -0.03 (-0.06:0.01), p=0.110 | -0.03 (-0.06:0.01), p=0.117 | -0.05 (-0.08:-0.01), p=0.009 |
| **Year of diagnosis (ASD)** | | 0.04 (0.01:0.07), p=0.005 | 0.01 (-0.02:0.04), p=0.655 | | | |
| **Age at diagnosis (ASD)** | | | 0.09 (0.06:0.12), p<0.001 | 0.09 (0.07:0.12), p<0.001 | 0.10 (0.06:0.13), p<0.001 | |
| **Year of birth** | | | | | 0.01 (-0.03:0.04), p=0.742 | -0.05 (-0.07:-0.02), p<0.001 |

**Supplementary Table 18. Linear regression models for Scizophrenia (SCZ) polygenic score in ASD patients N=17,071**. Abbreviations: PC=principal component; Year of diagnosis (ADHD)=first recorded ADHD diagnosis for each individuals between 1994:2016 obtained from Danish national registers; Age at diagnosis (ADHD)=age at first recorded ADHD diagnosis for each individual between 1:35 obtained from Danish national health registers; Year of birth= between 1981:2008 obtained from Danish national health registers.

| Predictor | Model 0 | Model 1 | Model 2 | Model 3 | Model 4 | Model 5 |
|---|---|---|---|---|---|---|
| **(Intercept)** | 0.06 (0.03:0.09), p<0.001 | 0.12 (0.07:0.18), p<0.001 | 0.10 (0.04:0.16), p=0.001 | 0.04 (-0.01:0.09), p=0.099 | 0.16 (0.07:0.25), p<0.001 | 0.12 (0.07:0.17), p<0.001 |
| **PC1** | 22.63 (17.62:27.64), p<0.001 | 22.60 (17.59:27.61), p<0.001 | 22.96 (17.94:27.98), p<0.001 | 22.79 (17.76:27.81), p<0.001 | 22.95 (17.93:27.97), p<0.001 | 23.07 (18.06:28.09), p<0.001 |
| **PC2** | 1.31 (-3.58:6.21), p=0.599 | 1.33 (-3.57:6.23), p=0.594 | 1.34 (-3.56:6.24), p=0.592 | 1.32 (-3.58:6.21), p=0.598 | 1.33 (-3.57:6.23), p=0.594 | 1.33 (-3.57:6.23), p=0.595 |
| **PC3** | -1.91 (-6.98:3.15), p=0.459 | -2.06 (-7.12:3.01), p=0.426 | -1.82 (-6.89:3.25), p=0.481 | -1.79 (-6.86:3.28), p=0.489 | -1.81 (-6.88:3.26), p=0.485 | -1.68 (-6.75:3.38), p=0.515 |
| **PC4** | -3.13 (-8.05:1.79), p=0.212 | -3.20 (-8.12:1.71), p=0.202 | -3.25 (-8.17:1.67), p=0.195 | -3.15 (-8.06:1.77), p=0.210 | -3.25 (-8.16:1.67), p=0.196 | -3.24 (-8.15:1.68), p=0.197 |
| **PC5** | -2.42 (-7.36:2.52), p=0.338 | -2.38 (-7.32:2.56), p=0.344 | -2.44 (-7.38:2.50), p=0.333 | -2.45 (-7.39:2.50), p=0.332 | -2.45 (-7.39:2.49), p=0.331 | -2.48 (-7.42:2.46), p=0.325 |
| **PC6** | 10.15 (5.21:15.10), p<0.001 | 10.17 (5.23:15.11), p<0.001 | 10.08 (5.13:15.02), p<0.001 | 10.11 (5.17:15.05), p<0.001 | 10.10 (5.15:15.04), p<0.001 | 10.06 (5.11:15.00), p<0.001 |
| **PC7** | 16.89 (12.00:21.79), p<0.001 | 16.87 (11.98:21.77), p<0.001 | 16.79 (11.90:21.69), p<0.001 | 16.86 (11.97:21.75), p<0.001 | 16.80 (11.91:21.69), p<0.001 | 16.78 (11.89:21.67), p<0.001 |
| **PC8** | 2.58 (-2.35:7.50), p=0.305 | 2.55 (-2.37:7.48), p=0.309 | 2.56 (-2.37:7.48), p=0.308 | 2.58 (-2.34:7.51), p=0.304 | 2.56 (-2.36:7.49), p=0.308 | 2.57 (-2.35:7.50), p=0.306 |
| **PC9** | -8.49 (-13.42:-3.57), p=0.001 | -8.52 (-13.44:-3.59), p=0.001 | -8.56 (-13.48:-3.64), p=0.001 | -8.51 (-13.44:-3.59), p=0.001 | -8.56 (-13.49:-3.64), p=0.001 | -8.57 (-13.49:-3.65), p=0.001 |
| **PC10** | -3.52 (-8.41:1.38), p=0.159 | -3.61 (-8.50:1.28), p=0.148 | -3.63 (-8.53:1.26), p=0.146 | -3.52 (-8.41:1.38), p=0.159 | -3.62 (-8.52:1.27), p=0.147 | -3.60 (-8.49:1.30), p=0.149 |
| **Sex** | 0.00 (-0.03:0.04), p=0.924 | -0.00 (-0.04:0.03), p=0.793 | -0.00 (-0.04:0.03), p=0.981 | 0.00 (-0.03:0.04), p=0.811 | -0.00 (-0.04:0.03), p=0.986 | 0.00 (-0.03:0.04), p=0.852 |
| **Year of diagnosis (ASD)** | | -0.04 (-0.07:-0.01), p=0.007 | -0.05 (-0.08:-0.02), p=0.001 | | | |
| **Age at diagnosis (ASD)** | | | 0.03 (0.00:0.06), p=0.047 | 0.01 (-0.01:0.04), p=0.353 | -0.02 (-0.05:0.01), p=0.245 | |
| **Year of birth** | | | | | -0.05 (-0.08:-0.02), p=0.002 | -0.04 (-0.06:-0.01), p=0.002 |

**Supplementary Table 19. Linear regression models for Addiction polygenic score in ASD patients N=17,071**. Abbreviations: PC=principal component; Year of diagnosis (ADHD)=first recorded ADHD diagnosis for each individuals between 1994:2016 obtained from Danish national registers; Age at diagnosis (ADHD)=age at first recorded ADHD diagnosis for each individual between 1:35 obtained from Danish national health registers; Year of birth= between 1981:2008 obtained from Danish national health registers.

| Predictor | Model 0 | Model 1 | Model 2 | Model 3 | Model 4 | Model 5 |
|---|---|---|---|---|---|---|
| **(Intercept)** | 0.06 (0.03:0.09), p<0.001 | 0.06 (0.01:0.12), p=0.027 | 0.03 (-0.03:0.09), p=0.385 | 0.01 (-0.04:0.05), p=0.806 | 0.05 (-0.04:0.14), p=0.311 | 0.11 (0.06:0.16), p<0.001 |
| **PC1** | 5.98 (0.95:11.01), p=0.020 | 5.98 (0.95:11.00), p=0.020 | 6.53 (1.49:11.57), p=0.011 | 6.47 (1.43:11.51), p=0.012 | 6.53 (1.49:11.56), p=0.011 | 6.35 (1.32:11.39), p=0.013 |
| **PC2** | -3.40 (-8.31:1.52), p=0.176 | -3.39 (-8.31:1.52), p=0.176 | -3.38 (-8.30:1.53), p=0.177 | -3.39 (-8.31:1.52), p=0.176 | -3.39 (-8.30:1.53), p=0.177 | -3.38 (-8.30:1.53), p=0.177 |
| **PC3** | -0.77 (-5.85:4.31), p=0.766 | -0.78 (-5.86:4.31), p=0.765 | -0.41 (-5.49:4.68), p=0.875 | -0.40 (-5.48:4.69), p=0.878 | -0.40 (-5.49:4.69), p=0.877 | -0.58 (-5.66:4.50), p=0.823 |
| **PC4** | -2.39 (-7.32:2.55), p=0.343 | -2.39 (-7.32:2.55), p=0.343 | -2.46 (-7.40:2.47), p=0.328 | -2.43 (-7.36:2.51), p=0.335 | -2.46 (-7.39:2.47), p=0.328 | -2.47 (-7.41:2.46), p=0.326 |
| **PC5** | -1.68 (-6.64:3.28), p=0.506 | -1.68 (-6.64:3.28), p=0.507 | -1.77 (-6.73:3.19), p=0.485 | -1.77 (-6.73:3.19), p=0.484 | -1.77 (-6.73:3.19), p=0.484 | -1.73 (-6.69:3.23), p=0.493 |
| **PC6** | 8.09 (3.13:13.05), p=0.001 | 8.09 (3.13:13.05), p=0.001 | 7.95 (2.99:12.91), p=0.002 | 7.96 (3.00:12.92), p=0.002 | 7.96 (3.00:12.92), p=0.002 | 8.01 (3.05:12.97), p=0.002 |
| **PC7** | 11.71 (6.80:16.62), p<0.001 | 11.71 (6.80:16.62), p<0.001 | 11.59 (6.68:16.50), p<0.001 | 11.61 (6.70:16.52), p<0.001 | 11.59 (6.68:16.50), p<0.001 | 11.62 (6.71:16.53), p<0.001 |
| **PC8** | 9.28 (4.33:14.22), p<0.001 | 9.28 (4.33:14.22), p<0.001 | 9.28 (4.34:14.23), p<0.001 | 9.29 (4.35:14.23), p<0.001 | 9.29 (4.35:14.23), p<0.001 | 9.27 (4.33:14.22), p<0.001 |
| **PC9** | 0.23 (-4.71:5.17), p=0.927 | 0.23 (-4.71:5.17), p=0.928 | 0.16 (-4.78:5.10), p=0.950 | 0.18 (-4.77:5.12), p=0.944 | 0.16 (-4.78:5.10), p=0.950 | 0.17 (-4.78:5.11), p=0.947 |
| **PC10** | 0.81 (-4.10:5.72), p=0.746 | 0.81 (-4.10:5.72), p=0.747 | 0.78 (-4.14:5.69), p=0.757 | 0.82 (-4.10:5.73), p=0.745 | 0.78 (-4.13:5.69), p=0.755 | 0.74 (-4.17:5.66), p=0.766 |
| **Sex** | -0.00 (-0.04:0.03), p=0.900 | -0.00 (-0.04:0.03), p=0.894 | 0.00 (-0.03:0.04), p=0.816 | 0.01 (-0.03:0.04), p=0.744 | 0.00 (-0.03:0.04), p=0.810 | -0.00 (-0.04:0.03), p=0.960 |
| **Year of diagnosis (ASD)** | | -0.00 (-0.03:0.03), p=0.944 | -0.02 (-0.05:0.01), p=0.257 | | | |
| **Age at diagnosis (ASD)** | | | 0.05 (0.02:0.07), p=0.002 | 0.04 (0.01:0.07), p=0.004 | 0.03 (-0.01:0.06), p=0.096 | |
| **Year of birth** | | | | | -0.02 (-0.05:0.01), p=0.297 | -0.03 (-0.06:-0.01), p=0.010 |

**Supplementary Table 20. Linear regression models for Education polygenic score in ASD patients N=17,071**. Abbreviations: PC=principal component; Year of diagnosis (ADHD)=first recorded ADHD diagnosis for each individuals between 1994:2016 obtained from Danish national registers; Age at diagnosis (ADHD)=age at first recorded ADHD diagnosis for each individual between 1:35 obtained from Danish national health registers; Year of birth= between 1981:2008 obtained from Danish national health registers.

| Predictor | Model 0 | Model 1 | Model 2 | Model 3 | Model 4 | Model 5 |
|---|---|---|---|---|---|---|
| **(Intercept)** | 0.06 (0.03:0.09), p<0.001 | 0.26 (0.21:0.32), p<0.001 | 0.32 (0.26:0.38), p<0.001 | 0.21 (0.16:0.26), p<0.001 | 0.45 (0.36:0.54), p<0.001 | 0.07 (0.02:0.11), p=0.009 |
| **PC1** | -26.34 (-31.38:-21.31), p<0.001 | -26.42 (-31.44:-21.40), p<0.001 | -27.36 (-32.39:-22.34), p<0.001 | -27.70 (-32.73:-22.66), p<0.001 | -27.36 (-32.39:-22.34), p<0.001 | -26.31 (-31.35:-21.27), p<0.001 |
| **PC2** | 1.26 (-3.65:6.18), p=0.614 | 1.32 (-3.59:6.22), p=0.599 | 1.30 (-3.61:6.20), p=0.604 | 1.25 (-3.66:6.16), p=0.617 | 1.28 (-3.62:6.19), p=0.608 | 1.26 (-3.65:6.18), p=0.614 |
| **PC3** | 4.64 (-0.45:9.72), p=0.074 | 4.18 (-0.89:9.26), p=0.106 | 3.55 (-1.52:8.63), p=0.170 | 3.61 (-1.47:8.69), p=0.164 | 3.58 (-1.49:8.66), p=0.167 | 4.66 (-0.43:9.74), p=0.073 |
| **PC4** | 1.15 (-3.79:6.09), p=0.649 | 0.93 (-4.00:5.86), p=0.712 | 1.06 (-3.87:5.98), p=0.074 | 1.26 (-3.67:6.19), p=0.617 | 1.06 (-3.87:5.98), p=0.074 | 1.14 (-3.80:6.08), p=0.651 |
| **PC5** | -0.08 (-5.05:4.88), p=0.973 | 0.02 (-4.93:4.97), p=0.994 | 0.17 (-4.78:5.12), p=0.947 | 0.16 (-4.79:5.11), p=0.950 | 0.15 (-4.80:5.09), p=0.954 | -0.09 (-5.05:4.87), p=0.972 |
| **PC6** | 1.69 (-3.28:6.65), p=0.505 | 1.74 (-3.21:6.69), p=0.491 | 1.99 (-2.96:6.94), p=0.432 | 2.05 (-2.90:7.01), p=0.417 | 2.02 (-2.93:6.97), p=0.424 | 1.68 (-3.28:6.64), p=0.507 |
| **PC7** | 1.82 (-3.09:6.74), p=0.467 | 1.76 (-3.14:6.66), p=0.482 | 1.97 (-2.93:6.87), p=0.430 | 2.10 (-2.81:7.01), p=0.402 | 1.98 (-2.92:6.88), p=0.428 | 1.81 (-3.10:6.73), p=0.470 |
| **PC8** | 5.19 (0.24:10.13), p=0.040 | 5.12 (0.18:10.05), p=0.042 | 5.10 (0.17:10.04), p=0.043 | 5.15 (0.21:10.08), p=0.041 | 5.11 (0.18:10.04), p=0.042 | 5.19 (0.24:10.13), p=0.040 |
| **PC9** | -11.75 (-16.69:-6.80), p<0.001 | -11.82 (-16.75:-6.88), p<0.001 | -11.70 (-16.63:-6.76), p<0.001 | -11.60 (-16.54:-6.67), p<0.001 | -11.71 (-16.64:-6.78), p<0.001 | -11.75 (-16.70:-6.81), p<0.001 |
| **PC10** | 4.15 (-0.77:9.07), p=0.098 | 3.87 (-1.04:8.77), p=0.122 | 3.92 (-0.98:8.82), p=0.117 | 4.14 (-0.77:9.04), p=0.098 | 3.93 (-0.97:8.83), p=0.116 | 4.14 (-0.77:9.06), p=0.098 |
| **Sex** | 0.03 (-0.00:0.07), p=0.088 | 0.01 (-0.02:0.05), p=0.562 | -0.00 (-0.04:0.03), p=0.961 | 0.01 (-0.03:0.04), p=0.650 | -0.00 (-0.04:0.03), p=0.956 | 0.03 (-0.00:0.07), p=0.087 |
| **Year of diagnosis (ASD)** | | -0.12 (-0.15:-0.10), p<0.001 | -0.10 (-0.13:-0.07), p<0.001 | | | |
| **Age at diagnosis (ASD)** | | | -0.08 (-0.11:-0.05), p<0.001 | -0.11 (-0.14:-0.08), p<0.001 | -0.17 (-0.21:-0.14), p<0.001 | |
| **Year of birth** | | | | | -0.10 (-0.13:-0.07), p<0.001 | -0.00 (-0.03:0.02), p=0.813 |

**Supplementary Table 21. Linear regression models for Intelligence polygenic score in ASD patients N=17,071**. Abbreviations: PC=principal component; Year of diagnosis (ADHD)=first recorded ADHD diagnosis for each individuals between 1994:2016 obtained from Danish national registers; Age at diagnosis (ADHD)=age at first recorded ADHD diagnosis for each individual between 1:35 obtained from Danish national health registers; Year of birth= between 1981:2008 obtained from Danish national health registers.

| Predictor | Model 0 | Model 1 | Model 2 | Model 3 | Model 4 | Model 5 |
|---|---|---|---|---|---|---|
| **(Intercept)** | 0.09 (0.06:0.12), p<0.001 | 0.15 (0.10:0.21), p<0.001 | 0.17 (0.11:0.23), p<0.001 | 0.13 (0.08:0.18), p<0.001 | 0.22 (0.13:0.31), p<0.001 | 0.10 (0.05:0.15), p<0.001 |
| **PC1** | -19.06 (-24.12:-14.00), p<0.001 | -19.08 (-24.14:-14.02), p<0.001 | -19.29 (-24.37:-14.22), p<0.001 | -19.41 (-24.48:-14.34), p<0.001 | -19.29 (-24.36:-14.22), p<0.001 | -19.98 (-24.04:-13.91), p<0.001 |
| **PC2** | 5.14 (0.19:10.08), p=0.042 | 5.15 (0.21:10.10), p=0.041 | 5.15 (0.20:10.09), p=0.041 | 5.13 (0.19:10.08), p=0.042 | 5.14 (0.20:10.09), p=0.041 | 5.14 (0.19:10.09), p=0.042 |
| **PC3** | 0.37 (-4.75:5.48), p=0.889 | 0.22 (-4.90:5.33), p=0.934 | 0.07 (-5.05:5.19), p=0.977 | 0.09 (-5.03:5.21), p=0.971 | 0.08 (-5.04:5.20), p=0.974 | 0.41 (-4.71:5.52), p=0.876 |
| **PC4** | 1.90 (-3.07:6.86), p=0.454 | 1.83 (-3.14:6.79), p=0.471 | 1.85 (-3.11:6.82), p=0.464 | 1.93 (-3.04:6.89), p=0.447 | 1.85 (-3.11:6.82), p=0.464 | 1.88 (-3.09:6.85), p=0.459 |
| **PC5** | -0.88 (-5.87:4.11), p=0.731 | -0.84 (-5.83:4.15), p=0.741 | -0.81 (-5.80:4.18), p=0.751 | -0.81 (-5.80:4.18), p=0.750 | -0.82 (-5.81:4.17), p=0.748 | -0.89 (-5.88:4.10), p=0.728 |
| **PC6** | 2.99 (-2.01:7.98), p=0.241 | 3.00 (-1.99:7.99), p=0.238 | 3.06 (-1.93:8.05), p=0.230 | 3.08 (-1.91:8.07), p=0.226 | 3.07 (-1.92:8.06), p=0.228 | 2.97 (-2.02:7.96), p=0.244 |
| **PC7** | -6.06 (-11.00:-1.12), p=0.016 | -6.08 (-11.02:-1.14), p=0.016 | -6.03 (-10.97:-1.09), p=0.017 | -5.99 (-10.93:-1.04), p=0.018 | -6.03 (-10.97:-1.09), p=0.017 | -6.08 (-11.02:-1.14), p=0.016 |
| **PC8** | 11.62 (6.64:16.59), p<0.001 | 11.60 (6.62:16.57), p<0.001 | 11.59 (6.62:16.57), p<0.001 | 11.61 (6.63:16.58), p<0.001 | 11.59 (6.62:16.57), p<0.001 | 11.62 (6.64:16.59), p<0.001 |
| **PC9** | -4.09 (-9.07:0.88), p=0.107 | -4.12 (-9.09:0.86), p=0.105 | -4.09 (-9.06:0.88), p=0.107 | -4.06 (-9.03:0.92), p=0.110 | -4.09 (-9.07:0.88), p=0.107 | -4.11 (-9.08:0.87), p=0.105 |
| **PC10** | 2.53 (-2.41:7.48), p=0.315 | 2.44 (-2.50:7.38), p=0.333 | 2.45 (-2.49:7.40), p=0.331 | 2.53 (-2.41:7.47), p=0.316 | 2.45 (-2.49:7.40), p=0.331 | 2.52 (-2.43:7.46), p=0.318 |
| **Sex** | 0.01 (-0.02:0.05), p=0.429 | 0.01 (-0.03:0.04), p=0.673 | 0.01 (-0.03:0.04), p=0.780 | 0.01 (-0.03:0.04), p=0.647 | 0.00 (-0.03:0.04), p=0.784 | 0.01 (-0.02:0.05), p=0.420 |
| **Year of diagnosis (ASD)** | | -0.04 (-0.07:-0.01), p=0.006 | -0.03 (-0.07:-0.00), p=0.030 | | | |
| **Age at diagnosis (ASD)** | | | -0.02 (-0.05:0.01), p=0.237 | -0.03 (-0.06:-0.00), p=0.039 | -0.05 (-0.09:-0.02), p=0.003 | |
| **Year of birth** | | | | | -0.04 (-0.07:-0.00), p=0.026 | -0.01 (-0.03:0.02), p=0.588 |

**Supplementary Table 22. Linear regression models for Risk-taking polygenic score in ASD patients N=17,071**. Abbreviations: PC=principal component; Year of diagnosis (ADHD)=first recorded ADHD diagnosis for each individuals between 1994:2016 obtained from Danish national registers; Age at diagnosis (ADHD)=age at first recorded ADHD diagnosis for each individual between 1:35 obtained from Danish national health registers; Year of birth= between 1981:2008 obtained from Danish national health registers.

| Predictor | Model 0 | Model 1 | Model 2 | Model 3 | Model 4 | Model 5 |
|---|---|---|---|---|---|---|
| **(Intercept)** | 0.02 (-0.01 : 0.05), 0.278 | 0.07 (0.02 : 0.13), 0.010 | 0.07 (0.01 : 0.13), 0.018 | 0.03 (-0.02 : 0.08), 0.194 | 0.12 (0.02 : 0.21), 0.013 | 0.04 (-0.01 : 0.09), 0.106 |
| **PC1** | 6.51 (1.41 : 11.61), 0.012 | 6.49 (1.39 : 11.59), 0.013 | 6.50 (1.39 : 11.61), 0.013 | 6.37 (1.26 : 11.48), 0.014 | 6.49 (1.38 : 11.60), 0.013 | 6.69 (1.58 : 11.80), 0.010 |
| **PC2** | 4.25 (-0.73 : 9.24), 0.094 | 4.27 (-0.72 : 9.25), 0.093 | 4.27 (-0.72 : 9.25), 0.093 | 4.25 (-0.73 : 9.24), 0.094 | 4.26 (-0.72 : 9.25), 0.094 | 4.26 (-0.73 : 9.24), 0.094 |
| **PC3** | -1.03 (-6.18 : 4.12), 0.695 | -1.16 (-6.31 : 3.99), 0.659 | -1.16 (-6.32 : 4.00), 0.660 | -1.14 (-6.30 : 4.03), 0.666 | -1.15 (-6.31 : 4.01), 0.664 | -0.94 (-6.09 : 4.22), 0.721 |
| **PC4** | -1.36 (-6.37 : 3.64), 0.593 | -1.42 (-6.43 : 3.58), 0.577 | -1.43 (-6.43 : 3.58), 0.577 | -1.35 (-6.36 : 3.65), 0.597 | -1.42 (-6.42 : 3.58), 0.578 | -1.40 (-6.41 : 3.60), 0.582 |
| **PC5** | 3.25 (-1.77 : 8.28), 0.205 | 3.28 (-1.74 : 8.31), 0.200 | 3.28 (-1.74 : 8.31), 0.201 | 3.28 (-1.75 : 8.31), 0.201 | 3.28 (-1.75 : 8.30), 0.202 | 3.23 (-1.80 : 8.26), 0.208 |
| **PC6** | 7.39 (2.36 : 12.42), 0.004 | 7.40 (2.37 : 12.43), 0.004 | 7.40 (2.37 : 12.43), 0.004 | 7.42 (2.39 : 12.46), 0.004 | 7.41 (2.38 : 12.44), 0.004 | 7.35 (2.32 : 12.38), 0.004 |
| **PC7** | -1.50 (-6.49 : 3.48), 0.554 | -1.52 (-6.50 : 3.46), 0.549 | -1.52 (-6.50 : 3.46), 0.549 | -1.48 (-6.46 : 3.50), 0.561 | -1.52 (-6.50 : 3.46), 0.551 | -1.55 (-6.53 : 3.43), 0.542 |
| **PC8** | -6.04 (-11.05 : -1.03), 0.018 | -6.06 (-11.07 : -1.05), 0.018 | -6.06 (-11.07 : -1.05), 0.018 | -6.05 (-11.06 : -1.03), 0.018 | -6.06 (-11.07 : -1.04), 0.018 | -6.04 (-11.06 : -1.03), 0.018 |
| **PC9** | -3.61 (-8.62 : 1.40), 0.158 | -3.63 (-8.64 : 1.38), 0.156 | -3.63 (-8.64 : 1.38), 0.156 | -3.60 (-8.61 : 1.42), 0.160 | -3.63 (-8.65 : 1.38), 0.155 | -3.64 (-8.66 : 1.37), 0.154 |
| **PC10** | -1.71 (-6.69 : 3.28), 0.502 | -1.79 (-6.77 : 3.20), 0.482 | -1.79 (-6.77 : 3.20), 0.482 | -1.71 (-6.69 : 3.28), 0.502 | -1.78 (-6.76 : 3.20), 0.484 | -1.74 (-6.72 : 3.25), 0.494 |
| **Sex** | 0.03 (-0.00 : 0.07), 0.092 | 0.02 (-0.01 : 0.06), 0.174 | 0.02 (-0.01 : 0.06), 0.176 | 0.03 (-0.01 : 0.06), 0.124 | 0.02 (-0.01 : 0.06), 0.173 | 0.03 (-0.00 : 0.07), 0.086 |
| **Year of diagnosis (ASD)** | | -0.04 (-0.06 : -0.01), 0.018 | -0.04 (-0.07 : -0.00), 0.026 | | | |
| **Age at diagnosis (ASD)** | | | 0.00 (-0.03 : 0.03), 0.979 | -0.01 (-0.04 : 0.02), 0.428 | -0.03 (-0.07 : -0.00), 0.058 | |
| **Year of birth** | | | | | -0.03 (-0.06 : -0.00), 0.036 | -0.02 (-0.04 : -0.01), 0.230 |

**Supplementary Table 23. Linear regression models for Neuroticism polygenic score in ASD patients N=17,071**. Abbreviations: PC=principal component; Year of diagnosis (ADHD)=first recorded ADHD diagnosis for each individuals between 1994:2016 obtained from Danish national registers; Age at diagnosis (ADHD)=age at first recorded ADHD diagnosis for each individual between 1:35 obtained from Danish national health registers; Year of birth= between 1981:2008 obtained from Danish national health registers.

| Predictor | Model 0 | Model 1 | Model 2 | Model 3 | Model 4 | Model 5 |
|---|---|---|---|---|---|---|
| **(Intercept)** | 0.14 (0.11 : 0.17), <0.001 | 0.08 (0.02 : 0.13), 0.006 | 0.03 (-0.03 : 0.09), 0.366 | 0.04 (-0.00 : 0.09), 0.076 | 0.01 (-0.08 : 0.10), 0.886 | 0.18 (0.13 : 0.23), <0.001 |
| **PC1** | 10.89 (5.86 : 15.91), <0.001 | 10.91 (5.89 : 15.94), <0.001 | 11.68 (6.65 : 16.72), <0.001 | 11.73 (6.70 : 16.76), <0.001 | 11.68 (6.65 : 16.72), <0.001 | 11.21 (6.18 : 16.24), <0.001 |
| **PC2** | -4.86 (-9.78 : 0.05), 0.052 | -4.88 (-9.79 : 0.03), 0.052 | -4.86 (-9.77 : 0.05), 0.052 | -4.86 (-9.77 : 0.05), 0.053 | -4.86 (-9.77 : 0.05), 0.052 | -4.85 (-9.77 : -0.06), 0.053 |
| **PC3** | -4.50 (-9.58 : -0.58), 0.083 | -4.36 (-9.44 : 0.72), 0.093 | -3.85 (-8.93 : 1.24), 0.138 | -3.86 (-8.94 : 1.23), 0.137 | -3.85 (-8.94 : 1.23), 0.138 | -4.33 (-9.41 : 0.75), 0.095 |
| **PC4** | -1.54 (-6.47 : 3.39), 0.540 | -1.48 (-6.41 : 3.46), 0.557 | -1.58 (-6.51 : 3.35), 0.530 | -1.61 (-6.54 : 3.32), 0.522 | -1.58 (-6.51 : 3.35), 0.530 | -1.62 (-6.55 : 3.32), 0.521 |
| **PC5** | 1.89 (-3.07 : 6.85), 0.455 | 1.86 (-3.10 : 6.82), 0.462 | 1.74 (-3.22 : 6.69), 0.492 | 1.74 (-3.21 : 6.69), 0.491 | 1.74 (-3.21 : 6.69), 0.491 | 1.85 (-3.11 : 6.80), 0.465 |
| **PC6** | -9.75 (-14.71 : -4.79), <0.001 | -9.76 (-14.72 : -4.80), <0.001 | -9.96 (-14.92 : -5.01), <0.001 | -9.97 (-14.93 : -5.02), <0.001 | -9.97 (-14.93 : -5.01), <0.001 | -9.82 (-14.78 : -4.86), <0.001 |
| **PC7** | -1.50 (-6.41 : 3.41), 0.550 | -1.48 (-6.39 : 3.43), 0.555 | -1.65 (-6.56 : 3.26), 0.509 | -1.67 (-6.58 : 3.24), 0.505 | -1.65 (-6.56 : 3.25), 0.509 | -1.58 (-6.49 : 3.33), 0.529 |
| **PC8** | -17.83 (-22.77 : -12.89), <0.001 | -17.81 (-22.75 : -12.87), <0.001 | -17.79 (-22.73 : -12.86), <0.001 | -17.80 (-22.74 : -12.86), <0.001 | -17.80 (-22.73 : -12.86), <0.001 | -17.83 (-22.77 : -12.89), <0.001 |
| **PC9** | 6.82 (1.88 : 11.76), 0.007 | 6.84 (1.90 : 11.78), 0.007 | 6.74 (1.80 : 11.68), 0.007 | 6.73 (1.79 : 11.66), 0.008 | 6.74 (1.80 : 11.68), 0.007 | 6.76 (1.82 : 11.70), 0.007 |
| **PC10** | 12.08 (7.17 : 16.99), <0.001 | 12.16 (7.25 : 17.07), <0.001 | 12.12 (7.21 : 17.03), <0.001 | 12.09 (7.18 : 16.99), <0.001 | 12.12 (7.21 : 17.03), <0.001 | 12.02 (7.11 : 16.93), <0.001 |
| **Sex** | -0.04 (-0.08 : -0.01), 0.014 | -0.04 (-0.07 : -0.00), 0.036 | -0.03 (-0.06 : 0.01), 0.116 | -0.03 (-0.06 : 0.01), 0.099 | -0.03 (-0.06 : 0.01), 0.117 | -0.04 (-0.08 : -0.01), 0.017 |
| **Year of diagnosis (ASD)** | | 0.04 (0.01 : 0.07), 0.012 | 0.01 (-0.02 : 0.04), 0.378 | | | |
| **Age at diagnosis (ASD)** | | | 0.06 (0.03 : 0.09), <0.001 | 0.07 (0.04 : 0.10), <0.001 | 0.08 (0.04 : 0.11), <0.001 | |
| **Year of birth** | | | | | 0.01 (-0.02 : 0.05), 0.344 | -0.03 (-0.05 : -0.00), 0.028 |

**Supplementary Table 24. Mutually adjusted linear regressions for five psychiatric polygenic scores (excl. iPSYCH) in ADHD patients N=20,111**. Abbreviations: PC=principal component; Year of diagnosis (ADHD)=first recorded ADHD diagnosis for each individuals between 1994:2016 obtained from Danish national registers; Age at diagnosis (ADHD)=age at first recorded ADHD diagnosis for each individual between 1:35 obtained from Danish national health registers; ADHD=attention deficit hyperactivity disorder; ASD=autism spectrum disorder; BP=bipolar disorder; MDD=major depressive disorder; SCZ=schizophrenia spectrum disorder

| Predictors | ADHD PGS (excl. iPSYCH) | ASD PGS (excl. iPSYCH) | BP PGS (excl. iPSYCH | MDD PGS (excl. iPSYCH) | SCZ PGS (excl.iPSYCH) |
|---|---|---|---|---|---|
| **(Intercept)** | 0.27 (0.20 : 0.33), <0.001 | 0.09 (0.02 : 0.15), 0.009 | 0.09 (0.02 : 0.15), 0.009 | 0.13 (0.06 : 0.19), <0.001 | 0.10 (0.03 : 0.17), 0.003 |
| **PC1** | 7.03 (2.36 : 11.70), 0.003 | 7.82 (3.12 : 12.51), 0.001 | 12.84 (8.05 : 17.63), <0.001 | 12.51 (7.87 : 17.14), <0.001 | 17.86 (13.07 : 22.66), <0.001 |
| **PC2** | -0.65 (-5.15 : 3.84), 0.776 | 7.70 (3.18 : 12.21), 0.001 | -1.31 (-5.91 : 3.30), 0.578 | -2.79 (-7.25 : 1.67), 0.220 | 1.28 (-3.33 : 5.89), 0.587 |
| **PC3** | -8.68 (-13.37 : -4.00), <0.001 | 0.27 (-4.44 : 4.98), 0.910 | -0.70 (-5.50 : 4.10), 0.775 | -5.23 (-9.89 : -0.58), 0.027 | -4.10 (-8.91 : 0.71), 0.095 |
| **PC4** | -0.83 (-5.35 : 3.68), 0.717 | 2.14 (-2.40 : 6.67), 0.356 | 2.61 (-2.01 : 7.24), 0.268 | 1.76 (-2.72 : 6.24), 0.442 | -2.62 (-7.25 : 2.02), 0.268 |
| **PC5** | 11.54 (7.03 : 16.06), <0.001 | 1.15 (-3.39 : 5.69), 0.620 | -0.93 (-5.56 : 3.70), 0.693 | 6.88 (2.39 : 11.36), 0.003 | -5.77 (-10.41 : -1.13), 0.015 |
| **PC6** | -1.52 (-6.01 : 2.98), 0.508 | -5.06 (-9.58 : -0.55), 0.028 | 1.21 (-3.40 : 5.82), 0.607 | -1.81 (-6.28 : 2.65), 0.426 | 7.97 (3.35 : 12.58), 0.001 |
| **PC7** | -7.56 (-12.06 : -3.05), 0.001 | -16.04 (-20.56 : -11.52), <0.001 | -6.89 (-11.51 : -2.28), 0.003 | -2.79 (-7.26 : 1.68), 0.221 | 12.86 (8.24 : 17.48), <0.001 |
| **PC8** | -10.54 (-15.04 : -6.05), <0.001 | -9.16 (-13.67 : -4.64), <0.001 | -6.34 (-10.95 : -1.73), 0.007 | -3.80 (-8.26 : 0.67), 0.096 | 0.56 (-4.05 : 5.18), 0.811 |
| **PC9** | 11.65 (7.11 : 16.18), <0.001 | 4.80 (0.24 : 9.35), 0.039 | -2.84 (-7.49 : 1.81), 0.231 | 3.03 (-1.47 : 7.53), 0.187 | -13.53 (-18.18 : -8.87), <0.001 |
| **PC10** | 1.89 (-2.64 : 6.42), 0.413 | 1.69 (-2.86 : 6.24), 0.467 | 1.55 (-3.09 : 6.20), 0.512 | 3.53 (-0.97 : 8.03), 0.124 | 0.32 (-4.34 : 4.97), 0.893 |
| **Sex** | 0.00 (-0.03 : 0.03), 0.814 | -0.02 (-0.05 : 0.01), 0.295 | -0.03 (-0.06 : 0.00), 0.074 | -0.05 (-0.08 : -0.02), 0.001 | -0.02 (-0.05 : 0.02), 0.350 |
| **Year of diagnosis (ASD)** | -0.03 (-0.05 : -0.01), 0.005 | -0.05 (-0.07 : -0.03), <0.001 | 0.06 (0.03 : 0.08), <0.001 | 0.12 (0.10 : 0.14), <0.001 | 0.05 (0.02 : 0.07), <0.001 |
| **Age at diagnosis (ASD)** | -0.00 (-0.04 : 0.03), 0.931 | 0.02 (-0.02 : 0.05), 0.307 | -0.04 (-0.08 : -0.00), 0.028 | -0.01 (-0.04 : 0.03), 0.698 | -0.05 (-0.09 : -0.02), 0.004 |

**Supplementary Table 25. Comparing linear and nonlinear effects of year and age in ADHD N=20,111.**

| | Model 1 (age) | | | | Model 2 (age adj) | | | | Model 3 (year) | | | |
|---|---|---|---|---|---|---|---|---|---|---|---|---|
| ***PGS*** | *R2_linear* | *P_linear* | *R2_nonlinear* | *P_nonlinear* | *R2_linear* | *P_linear* | *R2_nonlinear* | *P_nonlinear* | *R2_linear* | *P_linear* | *R2_nonlinear* | *P_nonlinear* |
| metaPRS_ADHD | 1.0E-04 | 1.6E-01 | 6.2E-05 | 5.4E-01 | 2.8E-06 | 8.1E-01 | 8.6E-05 | 4.2E-01 | 6.7E-04 | 2.4E-04 | 2.4E-05 | 7.8E-01 |
| metaPRS_ASD | 1.7E-03 | 7.1E-09 | 1.1E-03 | 1.9E-05 | 9.0E-04 | 2.0E-05 | 9.7E-04 | 5.1E-05 | 1.5E-03 | 5.9E-08 | 1.3E-04 | 2.7E-01 |
| metaPRS_BP | 1.2E-03 | 1.4E-06 | 4.9E-04 | 7.0E-03 | 1.6E-03 | 2.1E-08 | 4.8E-04 | 7.8E-03 | 1.0E-04 | 1.5E-01 | 2.0E-04 | 1.3E-01 |
| metaPRS_MDD | 7.4E-03 | 2.2E-34 | 8.3E-04 | 2.1E-04 | 7.0E-03 | 1.3E-32 | 8.2E-04 | 2.4E-04 | 4.5E-04 | 2.5E-03 | 4.4E-04 | 1.2E-02 |
| metaPRS_SCZ | 1.2E-03 | 6.3E-07 | 8.5E-04 | 1.7E-04 | 1.7E-03 | 4.1E-09 | 7.9E-04 | 3.4E-04 | 1.6E-04 | 7.4E-02 | 1.9E-04 | 1.5E-01 |
| pgs_addiction | 6.8E-03 | 9.6E-32 | 1.3E-03 | 1.5E-06 | 6.8E-03 | 5.8E-32 | 1.3E-03 | 1.2E-06 | 1.7E-04 | 6.7E-02 | 8.8E-05 | 4.1E-01 |
| pgs_EA | 3.6E-03 | 1.4E-17 | 1.1E-03 | 1.2E-05 | 2.8E-03 | 5.2E-14 | 1.1E-03 | 1.7E-05 | 9.3E-04 | 1.3E-05 | 3.5E-04 | 2.8E-02 |
| pgs_IQ | 6.7E-04 | 2.4E-04 | 4.0E-04 | 1.8E-02 | 5.7E-04 | 6.8E-04 | 4.0E-04 | 1.8E-02 | 1.0E-04 | 1.5E-01 | 2.3E-04 | 9.7E-02 |
| pgs_risk | 2.1E-04 | 3.9E-02 | 5.5E-04 | 3.9E-03 | 2.9E-04 | 1.7E-02 | 5.7E-04 | 3.2E-03 | 2.0E-05 | 5.2E-01 | 6.0E-05 | 5.5E-01 |
| pgs_neu | 3.3E-03 | 2.5E-16 | 1.5E-04 | 2.2E-01 | 3.0E-03 | 4.1E-15 | 1.5E-04 | 2.2E-01 | 2.7E-04 | 1.8E-02 | 1.8E-04 | 1.6E-01 |

**Abbreviations:**

| | |
|---|---|
| Model1 | pgs ~ age + covariates (10 first principal componenets + sex) |
| Model2 | pgs ~ age + year + covariates |
| Model3 | pgs ~ year + covariates |
| age | age of diagnosis |
| age adj | age of diagnosis adjusted for year of diagnosis |
| year | year of diagnosis |
| R2_linear | variance explained from anova comparing a model with covariates only to a model with covariate and variable(s) of interest |
| P_linear | p-value from anova of R2_linear |
| R2_nonlinear | variance explained from anova comparing a model with covariates and variable of interest to a model with covariates and three polynomials of the variable of interst (i.e., linear, quardratic, cubed) |
| P_nonlinear | p-value from R2_nonlinear |
| PGS | polygenic score |
| ADHD | attention deficit hyperactivity disorder |
| ASD | autism spectrum disorder |
| BP | bipolar disorder |
| MDD | major depressive disorder |

**Supplementary Table 26. Mutually adjusted linear regressions incl. sex interaction with year for 10 polygenic scores in ADHD patients N=20,111**. Abbreviations: PC=principal component; year ADHD=first recorded ADHD diagnosis for each individuals between 1994:2016 obtained from Danish national registers; age ADHD=age at first recorded ADHD diagnosis for each individual between age 1:35 obtained from Danish national health registers; PGS=polygenic score; ADHD=attention deficit hyperactivity disorder; ASD=autism spectrum disorder; BP=bipolar disorder; MDD=major depressive disorder; SCZ=schizophrenia spectrum disorder

| **Predictors** | **ADHD PGS** | **ASD PGS** | **BP PGS** | **MDD PGS** | **SCZ PGS** | **Addiction PGS** | **Education PGS** | **Intelligence PGS** | **Risk-taking PGS** | **Neuroticism PGS** |
|---|---|---|---|---|---|---|---|---|---|---|
| **(Intercept)** | 0.56 (-0.44:0.69), <0.001 | 0.32 (0.20:0.45), <0.001 | 0.10 (-0.03:0.23), 0.141 | 0.17 (0.05:0.30), 0.007 | 0.05 (-0.18:0.08), 0.433 | 0.20 (0.08:0.33), 0.002 | -0.15 (-0.27:0.02), 0.023 | 0.14 (-0.27:0.02), 0.024 | 0.22 (-0.03:0.22), 0.128 | 0.10 (-0.03:0.35), 0.128 |
| **PC1** | 8.01 (3.38:12.65), 0.001 | 11.48 (6.83:16.13), <0.001 | 11.83 (7.04:16.61), <0.001 | 12.73 (8.12:17.34), <0.001 | 19.72 (15.02:24.42), <0.001 | 8.73 (4.03:13.42), <0.001 | -27.90 (-32.51:-23.28), <0.001 | -20.51 (-23.14:-15.88), <0.001 | 3.64 (0.51:9.34), 0.129 | 5.77 (1.12:10.41), 0.015 |
| **PC2** | 2.00 (-2.46:6.46), 0.379 | 9.80 (5.33:14.27), <0.001 | -1.81 (-6.41:2.79), 0.441 | -2.25 (-6.69:2.18), 0.319 | 3.61 (-0.92:8.13), 0.118 | -3.38 (-7.90:1.13), 0.142 | 2.43 (-2.01:6.87), 0.283 | 0.43 (-4.02:4.88), 0.850 | 5.03 (0.51:9.55), 0.029 | -4.36 (-8.51:0.11), 0.056 |
| **PC3** | -7.82 (-12.47:-3.17), 0.001 | -6.46 (-11.12:-1.80), 0.007 | -1.69 (-6.49:3.11), 0.490 | -3.17 (-7.79:1.45), 0.179 | -3.88 (-8.59:0.84), 0.107 | -5.33 (-10.04:-0.62), 0.027 | -0.15 (-4.78:4.49), 0.950 | -1.15 (-5.79:3.49), 0.627 | -0.75 (-5.46:3.96), 0.756 | -4.85 (-9.51:-0.19), 0.041 |
| **PC4** | 3.45 (-1.03:7.93), 0.131 | -0.04 (-4.53:4.46), 0.988 | 3.40 (-1.22:8.02), 0.150 | 2.36 (-2.10:6.81), 0.300 | 0.40 (-4.14:4.94), 0.863 | -0.19 (-4.71:4.34), 0.934 | 2.33 (-6.79:2.13), 0.306 | -4.49 (-8.96:0.02), 0.049 | 0.36 (-4.18:4.90), 0.876 | 2.35 (-2.14:6.84), 0.305 |
| **PC5** | 1.48 (-3.00:5.96), 0.518 | -4.45 (-8.94:0.05), 0.052 | 0.89 (-3.74:5.51), 0.707 | 5.47 (1.02:9.93), 0.016 | -3.08 (-7.60:1.49), 0.187 | -0.89 (-5.43:3.66), 0.700 | -0.99 (-5.46:3.47), 0.663 | -6.13 (-10.61:-1.66), 0.007 | 1.02 (-3.52:5.56), 0.660 | 6.26 (1.77:10.75), 0.006 |
| **PC6** | -2.52 (-6.98:1.93), 0.267 | 4.27 (-0.20:8.75), 0.061 | 1.41 (-3.19:6.01), 0.548 | -2.92 (-7.36:1.51), 0.196 | 9.38 (4.86:13.90), <0.001 | 8.96 (4.44:13.47), <0.001 | 5.10 (0.65:9.55), 0.025 | 4.48 (0.03:8.94), 0.049 | 7.76 (3.24:12.28), 0.001 | -11.28 (-15.75:-6.81), <0.001 |
| **PC7** | 6.00 (1.54:10.47), 0.008 | -16.90 (-21.38:-12.42), <0.001 | -7.14 (-11.75:-2.53), 0.002 | -1.84 (-6.28:2.60), 0.417 | 14.38 (9.85:18.91), <0.001 | 9.41 (4.89:13.47), <0.001 | 1.67 (-2.78:6.12), 0.463 | -3.95 (-8.41:0.51), 0.083 | -2.59 (-7.12:1.93), 0.261 | -5.19 (-9.68:-0.71), 0.023 |
| **PC8** | -4.33 (-8.79:0.12), 0.057 | -3.53 (-8.01:0.94), 0.122 | -4.13 (-8.73:0.47), 0.079 | -7.43 (-11.86:-2.99), 0.001 | 1.95 (-2.58:6.47), 0.399 | 3.44 (-1.08:7.95), 0.136 | 4.10 (-0.34:8.55), 0.070 | 9.01 (4.55:13.46), <0.001 | -1.25 (-5.76:3.27), 0.589 | -21.37 (-25.84:-16.91), <0.001 |
| **PC9** | 19.47 (14.97:23.97), <0.001 | 7.56 (3.05:12.08), 0.001 | -4.64 (-9.28:0.00), 0.050 | 2.80 (-1.68:7.27), 0.220 | -10.64 (-15.21:-6.08), <0.001 | -4.48 (-9.04:0.08), 0.054 | -8.99 (-13.48:-4.51), <0.001 | -1.94 (-6.43:2.56), 0.398 | -3.95 (-8.51:0.61), 0.089 | 9.40 (4.89:13.91), <0.001 |
| **PC10** | 5.01 (0.52:9.50), 0.029 | 4.82 (0.31:9.33), 0.036 | 2.35 (-2.29:6.99), 0.321 | 3.34 (-1.13:7.80), 0.144 | -0.53 (-5.09:4.03), 0.821 | 0.56 (-3.99:5.12), 0.808 | 2.53 (-1.95:7.01), 0.268 | 0.83 (-3.66:5.32), 0.716 | -4.13 (-8.68:0.43), 0.076 | 4.20 (-0.31:8.70), 0.068 |
| **sex** | 0.03 (-0.11:0.17), 0.646 | -0.04 (-0.18:0.10), 0.557 | 0.02 (-0.12:0.17), 0.744 | -0.10 (-0.23:0.04), 0.169 | 0.07 (-0.07:0.21), 0.303 | -0.10 (-0.24:0.04), 0.147 | -0.12 (-0.26:0.01), 0.080 | -0.06 (-0.19:0.08), 0.430 | -0.07 (-0.21:0.07), 0.358 | -0.08 (-0.22:0.06), 0.259 |
| **age ADHD** | -0.00 (-0.02:0.02), 0.778 | -0.05 (-0.07:-0.03), <0.001 | 0.06 (0.04:0.09), <0.001 | 0.13 (0.11:0.15), <0.001 | 0.07 (0.04:0.09), <0.001 | 0.13 (0.11:0.15), <0.001 | -0.08 (-0.10:-0.06), <0.001 | -0.04 (-0.06:-0.02), 0.001 | 0.03 (0.00:0.05), 0.016 | 0.09 (0.07:0.11), <0.001 |
| **year ADHD** | -0.02 (-0.10:0.05), 0.509 | -0.06 (-0.14:0.01), 0.095 | -0.03 (-0.11:0.05), 0.433 | -0.04 (-0.11:0.04), 0.331 | -0.02 (-0.10:0.05), 0.561 | -0.08 (-0.16:-0.01), 0.026 | -0.09 (-0.16:-0.01), 0.019 | -0.04 (-0.11:0.04), 0.313 | -0.03 (-0.10:0.05), 0.461 | -0.03 (-0.11:0.04), 0.376 |
| **sex × year ADHD** | -0.04 (-0.13:0.04), 0.278 | -0.01 (-0.09:0.08), 0.901 | -0.03 (-0.12:0.05), 0.411 | 0.02 (-0.06:0.10), 0.568 | -0.05 (-0.14:0.03), 0.199 | 0.06 (-0.02:0.14), 0.158 | 0.07 (-0.01:0.15), 0.078 | 0.04 (-0.04:0.12), 0.312 | 0.00 (-0.08:0.09), 0.927 | 0.03 (-0.05:0.12), 0.404 |

**Supplementary Table 27. Mutually adjusted linear regressions for five psychiatric polygenic scores (excl. iPSYCH) in ASD patients N=17,071**. Abbreviations: PC=principal component; Year of diagnosis (ADHD)=first recorded ADHD diagnosis for each individuals between 1994:2016 obtained from Danish national registers; Age at diagnosis (ADHD)=age at first recorded ADHD diagnosis for each individual between 1:35 obtained from Danish national health registers; ADHD=attention deficit hyperactivity disorder; ASD=autism spectrum disorder; BP=bipolar disorder; MDD=major depressive disorder; SCZ=schizophrenia spectrum disorder

| **Predictors** | **ADHD PGS (excl. iPSYCH)** | **ASD PGS (excl. iPSYCH)** | **BP PGS (excl. iPSYCH** | **MDD PGS (excl. iPSYCH)** | **SCZ PGS (excl.iPSYCH)** |
|---|---|---|---|---|---|
| **(Intercept)** | 0.03 (-0.02 : 0.09), 0.250 | 0.26 (0.20 : 0.32), <0.001 | 0.14 (0.08 : 0.21), <0.001 | 0.14 (0.08 : 0.20), <0.001 | 0.20 (0.14 : 0.26), <0.001 |
| **PC1** | 5.72 (0.73 : 10.72), 0.025 | 12.18 (7.11 : 17.24), <0.001 | 15.73 (10.63 : 20.82), <0.001 | 14.78 (9.78 : 19.78), <0.001 | 19.61 (14.49 : 24.72), <0.001 |
| **PC2** | -7.80 (-12.67 : -2.93), 0.002 | 6.76 (1.81 : 11.70), 0.007 | -6.26 (-11.22 : -1.29), 0.014 | -1.48 (-6.36 : 3.40), 0.553 | -0.14 (-5.13 : 4.85), 0.955 |
| **PC3** | -7.98 (-13.02 : -2.94), 0.002 | 1.11 (-4.01 : 6.23), 0.671 | -3.02 (-8.16 : 2.12), 0.250 | -4.02 (-9.07 : 1.03), 0.119 | -2.43 (-7.59 : 2.74), 0.357 |
| **PC4** | -6.73 (-11.62 : -1.84), 0.007 | 1.24 (-3.72 : 6.21), 0.024 | 1.76 (-3.23 : 6.75), 0.489 | -0.11 (-5.01 : 4.79), 0.966 | -6.12 (-11.13 : -1.11), 0.017 |
| **PC5** | 4.51 (-0.40 : 9.42), 0.072 | 3.45 (-1.54 : 8.43), 0.176 | -0.49 (-5.50 : 4.52), 0.849 | 5.86 (0.94 : 10.79), 0.020 | -4.43 (-9.46 : 0.01), 0.085 |
| **PC6** | -1.36 (-6.27 : 3.56), 0.588 | -4.39 (-9.38 : 0.60), 0.085 | 5.34 (0.33 : 10.35), 0.037 | -3.26 (-8.18 : 1.66), 0.194 | 8.56 (3.52 : 13.59), 0.001 |
| **PC7** | -5.44 (-10.30 : -0.57), 0.028 | -15.90 (-20.84 : -10.96), <0.001 | 0.14 (-4.83 : 5.10), 0.957 | -3.61 (-8.48 : 1.27), 0.147 | 15.41 (10.43 : 20.40), <0.001 |
| **PC8** | -11.82 (-16.71 : -6.92), <0.001 | -11.15 (-16.13 : -6.18), <0.001 | -5.14 (-10.14 : -0.15), 0.044 | -2.05 (-6.95 : 2.86), 0.414 | 2.23 (-2.79 : 7.25), 0.384 |
| **PC9** | 17.27 (12.37 : 22.16), <0.001 | 1.10 (-3.87 : 6.07), 0.664 | -4.51 (-9.12 : 0.86), 0.105 | 0.23 (-4.68 : 5.14), 0.927 | -10.13 (-15.15 : -5.11), <0.001 |
| **PC10** | 3.58 (-1.28 : 8.45), 0.149 | 3.27 (-1.67 : 8.22), 0.194 | 2.21 (-2.75 : 7.17), 0.383 | 3.64 (-1.24 : 8.52), 0.144 | -1.80 (-6.79 : 3.19), 0.479 |
| **Sex** | 0.02 (-0.01 : 0.06), 0.238 | 0.05 (0.01 : 0.08), 0.011 | 0.02 (-0.01 : 0.06), 0.181 | -0.03 (-0.06 : 0.01), 0.158 | -0.00 (-0.04 : 0.03), 0.806 |
| **Year of diagnosis (ASD)** | -0.01 (-0.04 : 0.02), 0.375 | -0.07 (-0.10 : -0.04), <0.001 | 0.02 (-0.01 : 0.05), 0.287 | 0.07 (0.04 : 0.10), <0.001 | 0.02 (-0.01 : 0.05), 0.293 |
| **Age at diagnosis (ASD)** | 0.02 (-0.01 : 0.05), 0.153 | -0.04 (-0.07 : -0.01), 0.019 | -0.05 (-0.08 : -0.02), 0.001 | 0.00 (-0.03 : 0.03), 0.912 | -0.05 (-0.08 : -0.02), 0.002 |

**Supplementary Table 28. Comparing linear and nonlinear effects of year and age in ASD N=17,071.**

| | Model 1 (age) | | | | Model 2 (age adj) | | | | Model 3 (year) | | | |
|---|---|---|---|---|---|---|---|---|---|---|---|---|
| **PGS** | **R2_linear** | **P_linear** | **R2_nonlinear** | **P_nonlinear** | **R2_linear** | **P_linear** | **R2_nonlinear** | **P_nonlinear** | **R2_linear** | **P_linear** | **R2_nonlinear** | **P_nonlinear** |
| ADHD | 6.8E-05 | 2.8E-01 | 1.4E-03 | 4.2E-06 | 9.6E-08 | 9.7E-01 | 1.3E-03 | 1.1E-05 | 5.3E-04 | 2.5E-03 | 3.7E-04 | 4.3E-02 |
| ASD | 3.6E-04 | 1.3E-02 | 1.2E-05 | 9.0E-01 | 3.2E-05 | 4.6E-01 | 1.1E-06 | 9.9E-01 | 1.6E-03 | 2.4E-07 | 2.3E-04 | 1.4E-01 |
| BP | 2.1E-05 | 5.5E-01 | 7.8E-04 | 1.2E-03 | 1.7E-04 | 8.9E-02 | 6.6E-04 | 3.4E-03 | 4.9E-04 | 3.9E-03 | 5.2E-05 | 6.4E-01 |
| MDD | 2.8E-03 | 4.5E-12 | 3.5E-04 | 4.8E-02 | 2.3E-03 | 2.3E-10 | 3.5E-04 | 4.8E-02 | 4.6E-04 | 4.9E-03 | 1.2E-05 | 9.0E-01 |
| SCZ | 5.0E-05 | 3.5E-01 | 8.9E-04 | 4.7E-04 | 2.3E-04 | 4.7E-02 | 7.7E-04 | 1.3E-03 | 4.3E-04 | 6.7E-03 | 3.4E-04 | 5.5E-02 |
| Addiction | 4.8E-04 | 4.1E-03 | 2.9E-05 | 7.8E-01 | 5.6E-04 | 2.0E-03 | 3.0E-05 | 7.7E-01 | 2.8E-07 | 9.4E-01 | 1.2E-04 | 3.4E-01 |
| Education | 3.6E-03 | 3.7E-15 | 2.5E-03 | 2.8E-10 | 1.6E-03 | 1.4E-07 | 2.1E-03 | 1.0E-08 | 4.2E-03 | 2.1E-17 | 1.9E-04 | 2.0E-01 |
| Intelligence | 2.5E-04 | 3.9E-02 | 1.1E-04 | 3.9E-01 | 8.2E-05 | 2.4E-01 | 8.1E-05 | 5.0E-01 | 4.4E-04 | 6.0E-03 | 8.8E-05 | 4.7E-01 |
| Risk-taking | 3.7E-05 | 4.3E-01 | 6.4E-05 | 5.8E-01 | 4.0E-08 | 9.8E-01 | 7.0E-05 | 5.5E-01 | 3.3E-04 | 1.8E-02 | 1.6E-04 | 2.5E-01 |
| Neuroticism | 1.4E-03 | 9.4E-07 | 1.1E-04 | 4.0E-01 | 1.1E-03 | 1.7E-05 | 1.0E-04 | 4.2E-01 | 3.7E-04 | 1.2E-02 | 2.9E-05 | 7.8E-01 |

**Abbreviations:**

| | |
|---|---|
| Model1 | pgs ~ age + covariates (10 first principal componenets + sex) |
| Model2 | pgs ~ age + year + covariates |
| Model3 | pgs ~ year + covariates |
| age | age of diagnosis |
| age adj | age of diagnosis adjusted for year of diagnosis |
| aear | year of diagnosis |
| R2_linear | variance explained from anova comparing a model with covariates only to a model with covariate and variable(s) of interest |
| P_linear | p-value from anova of R2_linear |
| R2_nonlinear | variance explained from anova comparing a model with covariates and variable of interest to a model with covariates and three polynomials of the variable of interst (i.e., linear, quardratic, cubed) |
| P_nonlinear | p-value from R2_nonlinear |
| PGS | polygenic score |
| ADHD | attention deficit hyperactivity disorder |
| ASD | autism spectrum disorder |
| BP | bipolar disorder |
| MDD | major depressive disorder |

**Supplementary Table 29. Mutually adjusted linear regressions incl. sex interaction with year for 10 polygenic scores in ASD patients N=17,071**. Abbreviations: PC=principal component; year ASD=first recorded ASD diagnosis for each individual between 1994:2016 obtained from Danish national registers; age ASD=age at first recorded ASD diagnosis for each individual between age 1:35 obtained from Danish national health registers; PGS=polygenic score; ADHD=attention deficit hyperactivity disorder; ASD=autism spectrum disorder; BP=bipolar disorder; MDD=major depressive disorder; SCZ=schizophrenia spectrum disorder

| Predictor | ADHD PGS | ASD PGS | BP PGS | MDD PGS | SCZ PGS | Addiction PGS | Education PGS | Intelligence PGS | Risk-taking PGS | Neuroticism PGS |
|---|---|---|---|---|---|---|---|---|---|---|
| **Intercept** | -3.42 (-6.80:-0.04), 0.009 | 0.35 (0.24:0.46), <0.001 | 0.06 (-0.01:0.21), 0.069 | -0.10 (-0.31:0.11), 0.952 | 0.06 (-0.04:0.17), 0.247 | 0.02 (-0.09:0.13), 0.797 | 0.31 (0.20:0.42), <0.001 | 0.12 (-0.01:0.23), 0.032 | 0.04 (-0.07:0.15), 0.454 | 0.03 (-0.08:0.13), 0.646 |
| **PC1** | 13.04 (8.00:18.08), <0.001 | 11.91 (6.87:16.96), <0.001 | 14.12 (9.03:19.22), <0.001 | 15.30 (10.32:20.29), <0.001 | 22.93 (17.91:27.96), <0.001 | 6.52 (1.49:11.56), 0.011 | -27.37 (-32.40:-22.34), <0.001 | -19.33 (-24.49:-14.26), <0.001 | 6.47 (1.36:11.59), 0.013 | 11.68 (6.65:16.72), <0.001 |
| **PC2** | -2.71 (-7.62:2.21), 0.281 | 8.23 (3.31:13.15), 0.001 | -6.39 (-11.35:-1.42), 0.012 | -1.19 (-5.96:3.76), 0.658 | 1.33 (-3.56:6.23), 0.593 | -3.38 (-8.30:1.53), 0.177 | 1.30 (-3.61:6.20), 0.605 | 5.14 (0.29:10.09), 0.042 | 4.26 (-0.72:9.25), 0.094 | -4.86 (-9.77:0.05), 0.052 |
| **PC3** | -8.32 (-13.41:-3.24), 0.001 | -7.57 (-12.66:-2.47), 0.004 | -5.25 (-10.39:-0.11), 0.045 | -1.95 (-6.98:3.09), 0.448 | -1.83 (-6.90:3.24), 0.479 | -0.41 (-5.50:4.68), 0.874 | 3.55 (-1.53:8.63), 0.170 | 0.06 (-5.06:5.18), 0.981 | -1.16 (-6.32:4.00), 0.658 | -1.85 (-6.93:1.24), 0.138 |
| **PC4** | 3.88 (-1.06:8.81), 0.124 | 2.76 (-2.18:7.70), 0.273 | 4.31 (-0.67:9.30), 0.090 | 0.71 (-4.17:5.59), 0.776 | -3.25 (-8.16:1.67), 0.195 | -2.46 (-7.40:2.47), 0.328 | 1.06 (-3.87:5.98), 0.674 | 1.86 (-3.11:6.82), 0.463 | -1.42 (-6.43:3.58), 0.577 | -1.58 (-6.51:3.35), 0.530 |
| **PC5** | 0.90 (-4.05:5.86), 0.723 | -3.26 (-8.23:1.70), 0.197 | 3.88 (-1.13:8.89), 0.129 | 4.05 (-0.86:8.95), 0.106 | -2.45 (-7.35:2.49), 0.330 | -1.77 (-6.73:3.19), 0.484 | 0.16 (-4.78:5.11), 0.948 | -0.83 (-5.82:4.16), 0.743 | 2.27 (-1.76:6.30), 0.202 | 1.74 (-3.22:6.69), 0.492 |
| **PC6** | -0.39 (-5.35:4.58), 0.879 | 3.49 (0.51:6.46), 0.030 | 5.63 (0.62:10.64), 0.028 | -3.16 (-8.07:1.75), 0.207 | 10.99 (5.14:15.93), <0.001 | 7.95 (2.99:12.91), 0.002 | 1.99 (-2.96:6.94), 0.431 | 3.07 (-1.92:8.06), 0.228 | 7.41 (2.38:12.44), 0.004 | -9.96 (-14.82:-5.11), <0.001 |
| **PC7** | 8.64 (3.72:13.55), 0.001 | -9.58 (-14.50:-4.65), <0.001 | -4.52 (-5.48:-4.44), 0.837 | -1.71 (-6.57:3.15), 0.490 | 16.80 (11.91:21.69), <0.001 | 11.59 (6.68:16.50), <0.001 | 1.97 (-2.93:6.87), 0.430 | -6.02 (-10.96:-1.08), 0.017 | -1.52 (-6.50:3.46), 0.551 | -1.65 (-6.56:3.26), 0.510 |
| **PC8** | -5.12 (-10.06:-0.18), 0.042 | -4.32 (-9.27:-0.63), 0.087 | -3.77 (-8.76:1.23), 0.139 | -6.35 (-11.24:-1.45), 0.011 | 2.56 (-2.36:7.49), 0.308 | 9.29 (4.34:14.23), <0.001 | 5.11 (0.17:10.04), 0.042 | 11.60 (6.62:16.57), <0.001 | -0.06 (-11.07:3.95), 0.018 | -17.79 (-22.73:-12.85), <0.001 |
| **PC9** | 17.94 (13.00:22.89), <0.001 | 7.66 (2.71:12.61), 0.002 | -5.66 (-10.66:-0.67), 0.026 | 0.35 (-4.54:5.24), 0.890 | -8.56 (-13.49:-3.64), 0.001 | 0.16 (-4.78:5.19), 0.350 | -11.70 (-16.63:-6.77), <0.001 | -4.09 (-9.07:0.88), 0.107 | -3.63 (-8.65:1.38), 0.155 | 6.74 (1.80:11.68), 0.007 |
| **PC10** | 6.59 (1.67:11.50), 0.009 | 5.27 (0.35:10.19), 0.036 | 3.63 (-1.33:8.60), 0.151 | 3.93 (-0.93:8.79), 0.113 | -3.65 (-8.54:1.25), 0.144 | 0.77 (-4.14:5.68), 0.758 | 3.92 (-0.99:8.82), 0.117 | 2.43 (-2.51:7.37), 0.336 | 1.80 (-0.78:3.18), 0.479 | 12.12 (7.21:17.02), <0.001 |
| **gender** | 0.18 (0.06:0.29), 0.003 | 0.14 (0.02:0.26), 0.019 | 0.11 (-0.01:0.23), 0.072 | 0.12 (-0.00:0.23), 0.050 | 0.04 (-0.07:0.16), 0.473 | 0.02 (-0.10:0.13), 0.791 | 0.01 (-0.11:0.13), 0.836 | 0.07 (-0.05:0.18), 0.272 | 0.06 (-0.06:0.18), 0.297 | -0.03 (-0.14:0.09), 0.669 |
| **age ASD** | -0.00 (-0.03:0.03), 0.954 | -0.01 (-0.04:0.02), 0.409 | 0.02 (-0.00:0.05), 0.104 | 0.09 (0.06:0.12), <0.001 | 0.03 (-0.00:0.06), 0.051 | 0.05 (0.02:0.07), 0.002 | -0.08 (-0.11:-0.05), <0.001 | -0.02 (-0.05:0.01), 0.219 | -0.00 (-0.03:0.03), 0.997 | 0.06 (0.03:0.09), <0.001 |
| **year ASD** | 0.11 (0.04:0.17), 0.001 | -0.02 (-0.08:0.05), 0.634 | -0.01 (-0.07:0.06), 0.874 | 0.08 (0.02:0.15), 0.014 | -0.03 (-0.09:0.04), 0.393 | -0.01 (-0.08:0.05), 0.724 | -0.09 (-0.15:-0.02), 0.007 | -0.00 (-0.07:0.06), 0.933 | -0.02 (-0.08:0.05), 0.645 | 0.02 (-0.05:0.08), 0.647 |
| **gender × year ASD** | -0.08 (-0.15:-0.01), 0.031 | -0.07 (-0.14:0.00), 0.056 | -0.06 (-0.13:0.01), 0.113 | -0.09 (-0.16:-0.02), 0.011 | -0.03 (-0.10:0.04), 0.448 | -0.01 (-0.08:0.06), 0.838 | -0.01 (-0.08:0.06), 0.816 | -0.04 (-0.11:0.03), 0.287 | -0.02 (-0.10:0.05), 0.505 | -0.00 (-0.07:0.07), 0.962 |

**Supplementary Table 30.** iPSYCH Study Consortium banner authors.

| Group | First Names | Last Names | Email | Affiliations | Conflicts | ORCID | Grants |
|---|---|---|---|---|---|---|---|
| iPSYCH PI | Merete | Nordentoft | merete.nordentc | Mental Health Centre Copenhagen, Capital Region of Denmark, Copenhagen University Hospital, Copenhagen, Denmark | | | |
| iPSYCH PI | Ole Mors | Mors | | Psychosis Research Unit, Aarhus University Hospital-Psychiatry, Denmark | | | |
| iPSYCH PI | Preben Bo | Mortensen | | NCRR - National ( | None | | |
| Aarhus | Ditte | Demontis | | Center for Genomics and Personalized Medicine, Aarhus, Denmark | | | |
| Aarhus | Jakob | Grove | grove@biomed.a | Centre for Integrative Sequencing, Department of Biomedicine and iSEQ, Aarhus University, Aarhus, Denmark; BiRC Bioinformatics Research Centre, Aarhus University, Aarhus, Denmark | | | |
| Aarhus | Anna | Starnawska |.d | Department of Biomedicine, Aarhus University, Aarhus, Denmark | | | |
| Aarhus | Thomas | Damm Als | thomas.als@clin. | Centre for Integrative Sequencing, Department of Biomedicine and iSEQ, Aarhus University, Aarhus, Denmark | | | |
| IBP | Alfonso | Buil | alfonso.buil.dem | Institute of Biolo | None | | |
| IBP | Anders | Rosengren | anders.rosengrer | Institute of Biolo | None | 0000-0002-6682-1288 | |
| IBP | Andres | Ingasson | andres.ingason@ | Institute of Biolo | None | | |
| IBP | Dorte | Helenius | dorte.helenius.m | Institute of Biolo | None | | |
| IBP | Richard | Zetterberg | richard.zetterber | Institute of Biolo | None | | |
| NCRR | Carsten | Bøcker Pedersen | | National Centre for Register-based Research, BSS, Aarhus University | | | |
| NCRR | Jakob | Christensen |.d | National Centre for Register-based Research, BSS, Aarhus University | | | |
| NCRR | Liselotte | Pedersen | | National Centre for Register-based Research, BSS, Aarhus University | | | |
| NCRR | Marianne | Giørtz Pedersen | | National Centre for Register-based Research, BSS, Aarhus University | | | |
| SSI | Jonas | Bybjerg-Grauhol | | Department for ( | None | 0000-0003-1705-4008 | |
| SSI | Marie | Bækvad-Hansen | | Department for ( | None | 0000-0002-5881-1776 | |
